# Image-based & machine learning-guided multiplexed serology test for SARS-CoV-2

**DOI:** 10.1101/2022.09.08.22279729

**Authors:** Vilja Pietiäinen, Minttu Polso, Ede Migh, Christian Guckelsberger, Maria Harmati, Akos Diosdi, Laura Turunen, Antti Hassinen, Swapnil Potdar, Annika Koponen, Edina Gyukity Sebestyen, Ferenc Kovacs, Andras Kriston, Reka Hollandi, Katalin Burian, Gabriella Terhes, Adam Visnyovszki, Eszter Fodor, Zsombor Lacza, Anu Kantele, Pekka Kolehmainen, Laura Kakkola, Tomas Strandin, Lev Levanov, Olli Kallioniemi, Lajos Kemeny, Ilkka Julkunen, Olli Vapalahti, Krisztina Buzas, Lassi Paavolainen, Peter Horvath, Jussi Hepojoki

## Abstract

Here, we describe a scalable and automated, high-content microscopy -based mini-immunofluorescence assay (mini-IFA) for serological testing i.e., detection of antibodies. Unlike conventional IFA, which often relies on the use of cells infected with the target pathogen, our assay employs transfected cells expressing individual viral antigens. The assay builds on a custom neural network-based image analysis pipeline for the automated and multiplexed detection of immunoglobulins (IgG, IgA, and IgM) in patient samples. As a proof-of-concept, we employed high-throughput equipment to set up the assay for measuring antibody response against severe acute respiratory syndrome coronavirus 2 (SARS-CoV-2) infection with spike (S), membrane (M), and nucleo (N) proteins, and the receptor-binding domain (R) as the antigens. We compared the automated mini-IFA results from hundreds of patient samples to the visual observations of human experts and to the results obtained with conventional ELISA. The comparisons demonstrated a high correlation to both, suggesting high sensitivity and specificity of the mini-IFA. By testing pre-pandemic samples and those collected from patients with RT-PCR confirmed SARS-CoV-2 infection, we found mini-IFA to be most suitable for IgG and IgA detection. The results demonstrated N and S proteins as the ideal antigens, and the use of these antigens can serve to distinguish between vaccinated and infected individuals. The assay principle described enables detection of antibodies against practically any pathogen, and none of the assay steps require high biosafety level environment. The simultaneous detection of multiple Ig classes allows for distinguishing between recent and past infection.

**Public abstract:** The manuscript describes a miniaturized immunofluorescence assay (mini-IFA) for measuring antibody response in patient blood samples. The automated method builds on machine-learning -guided image analysis with SARS-CoV-2 as the model pathogen. The method enables simultaneous measurement of IgM, IgA, and IgG responses against different virus antigens in a high throughput manner. The assay relies on antigens expressed through transfection and allows for differentiation between vaccine-induced and infection-induced antibody responses. The transfection-based antigen expression enables performing the assay at a low biosafety level laboratory and allows fast adaptation of the assay to emerging pathogens. Our results provide proof-of-concept for the approach, demonstrating fast and accurate measurement of antibody responses in a clinical and research set-up.

## Main text

Serological assays are essential for studying and controlling infectious disease outbreaks, such as the current COVID-19 (coronavirus disease 2019) pandemic caused by severe acute respiratory syndrome coronavirus 2 (SARS-CoV-2). Immunofluorescence assays relying on cells infected with a given virus provide rapidly available means for demonstrating antibody response against emerging pathogens. However, setting up such assays may require a high biosafety level facility for handling the virus^1^. In addition, such assays are often low-throughput and interpretation of the results is labor-intensive and subjective. Here, we describe a mini-immunofluorescence assay (mini-IFA), which is a machine learning-guided, microscopy-based, automated, high-throughput serology assay, requiring a low biosafety level environment, and demonstrate its’ efficacy using SARS-CoV-2 as a model (**Fig. 1**).

**Figure 1.**
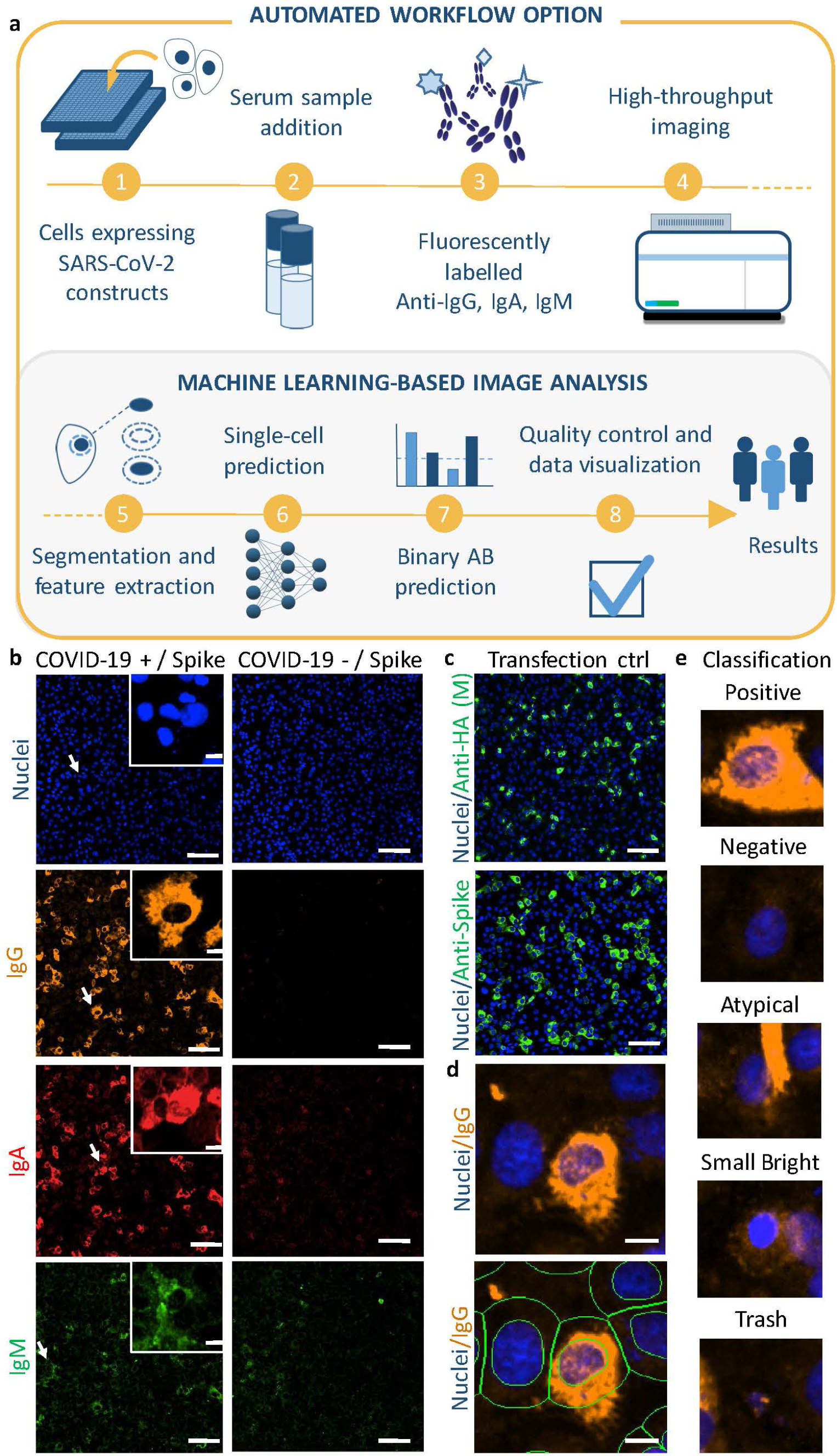
**a)** Assay workflow. Shortly, Vero E6 cells, transfected to express SARS-CoV-2 antigens (N, M, R, and S), are fixed in 384-well plates for incubation with serum samples. The immunoglobulins (IgG, IgA, and IgM) are detected simultaneously with fluorophore-labelled secondary antibodies using automated high-content fluorescence microscopy. Machine learning is employed for (i) nuclei and cell segmentation, and (ii) phenotypic cell classification. The results are presented via a decision-support system, which also allows interactive visual observation of the raw images. For a full explanation with numbers, see the main text. **b)** Examples of microscopy images of specific IgG (DL550, orange), IgA (AF647, red), and IgM (AF488, green) responses against SARS-CoV-2 spike (S) protein in assay control serum samples obtained from positive (COVID-19+) and negative (COVID-19–) patients. (Scale bar in overview images: 100 μm; zoom-in images: 10 μm.) **c)** Transfection efficiency of viral antigens is determined by immunostaining. Here, AF488-conjugated anti-HA (green) antibody for HA-tagged M protein and in-house rabbit anti-S antibody (AF488, green) for S protein are shown. Scale bars: 100 μm. **d)** Visualization of non-segmented and segmented cells. After segmentation of cell nuclei, an additional mask is created for the ‘whole cell’ by dilating the nucleus area for a maximum of 7 μm, and the ‘cytoplasm’ is determined as an area without the segmented nucleus. (Scale bars: 10 μm). **e)** Examples of manual labelling of cell classes for the training set. Cells were divided into five categories: positive, negative, atypical, small bright and trash. Unless otherwise stated, the microscopy images presented show cells expressing the S protein of SARS-CoV-2, and they have been linearly adjusted from the original 16-bit image format to improve visual appearance.

## Method

### Assay set-up and automation

#### Laboratory method

(Steps 1–4, **Fig. 1a**; **Supp. Text File 1** for the standard operating procedure). The cell line of interest (here: African green monkey kidney cells, Vero E6) is separately transfected with plasmids encoding four SARS-CoV-2 antigens (spike protein [S] with a His-tag; S-pCAGGS^2^, membrane protein [M] with an HA-tag; M-pEBB, nucleoprotein [N] with a His-tag; NP-pCAGGS^3^, and the receptor-binding domain, RBD [R], of S protein with His-tag; RBD-pCAGGS^2^) (see **Supp. Text File 2** for Materials & Methods, **and Table S1**). The transfected cells in suspension are transferred to 384-well assay plates with high-throughput automation. Transient transfection, as opposed to stable transfection, allows simultaneous control of the background signal and detection of autoantibodies, based on non-transfected cells included in the same well. Patient sera and assay controls (**Fig. 1b; Fig. S1**), as well as a DNA binding dye (Hoechst 33342), are added to the wells with an acoustic dispenser to simultaneously monitor the accuracy and success of the sample transfer. Each of assays (and plates) includes several positive (and negative) controls, which allow for comparing the results between assays and the measurement of transfection percentage, enabling cross-assay standardization. Immunoglobulin (Ig) classes present in the samples are detected by multiplexed immunostaining with fluorescent-labelled antibodies for IgM (AlexaFluor; AF; AF488), IgG (DyLight; DL; DL550), and IgA (AF647). Transfection efficiency (**Table S2**) is controlled by staining with antibodies (**Table S1**) targeting the respective (His/HA -tagged) antigens and/or antigen specific antisera. We image four different fluorescent channels with high-content microscopy to detect a) *IgG, b) IgA, c), IgM*) and d) the cell nuclei (**Fig. 1b-c; Fig. S1**). The method can be modified to include any antibodies/markers; however, any new marker will require a new training set for machine-learning models (see later assay adaptability & semi-automated method). Here, we excluded the cytoplasmic marker to include the transfection controls/additional Ig classes to the basic four channel microscopy set-up.

#### Computational Analysis (Steps 5-8, Fig. 1A)

The analysis pipeline includes the processing of the microscopy images, per-cell and per-well predictions, and the visualization of the results. The images were processed with the BIAS software^4,5^, facilitating pre-processing^6^, deep learning-based cell segmentation^7^ (**Fig. 1d**), and feature extraction (**Supp. Text File 2**). We developed dedicated Python scripts to use these features for the prediction of antibody response through supervised machine-learning (GitHub: https://github.com/fimm-covid-19-hca/mini-IFA_paper). More specifically, we trained models to classify each segmented immunostaining phenotype - represented by the extracted image features - into five classes: “positive”, “negative”, “atypical”, “small bright”, and “trash” **(Fig. 1e, Fig. S2; class design rationale in Supp. Text File 2)**. As training data, we labelled 55496 cells across 16 plates (four plates for each antigen), with roughly equal proportions per antigen **(Table S3)**. Labels were assigned in BIAS software^4,5^ as previously described^8^, using an active learning feature^9^ to increase training data quality. We avoid model bias through class imbalances by including class weights in the training. To account for potential signal variations across plates and batches, image features were normalized based on per-plate controls. We employed leave-one-plate-out cross-validation to (i) determine the best out of three normalization approaches and (ii) select the best classification model and hyperparameters for each Ig class-antigen pair. Performance was scored on prediction sensitivity **x** specificity, and one plate was left out as validation set in each fold to assess the method’s generalization potential as prediction quality on unseen plates – a typical scenario for the assay’s clinical application. We found that the model can classify individual cells accurately with specificity 0.96-0.97, 0.95-0.96, 0.96-0.97 and sensitivity 0.84-0.89, 0.79-0.84, 0.82-0.86, for IgG, IgA and IgM, respectively (**Supp. Text File 2, Fig. S3a-b, and Table S4**). Following this selection, we re-trained the best classification model/hyperparameter configuration on the entire training dataset, processed with the best normalization scheme. We applied the trained model to a separate experimental test dataset including samples from four plates, separately for each antigen to produce per-cell predictions. Based on these predictions, we calculated a per-well positivity score as the ratio of predicted positive cells over all segmented cells. The user can define the cut-off of this value based on positive and negative controls (See **Supp. Data 1** for data visualization graphs showing the sample distribution as compared to the positive and negative controls), resulting in binary per-well antibody response predictions.

#### Visualization

Quality control (QC) assessment with an automated script is available to control the assay’s quality, including cell seeding and sample transfer accuracy, as well as signal distribution for positive and negative controls (**Fig. S4)**. The data are visualized with (interactive) plots and heatmaps (**Supp. Data 1-2**).

#### Adaptability and scalability of the assay

To 1) evaluate the assay’s technical scalability and adaptability to a laboratory without advanced automation and to 2) test a variation that measures the IgG/IgA signal only in antigen transfected cells (recognized with his-tag), either shipped ready-to-go assay plates (cells transfected with the virus antigen and fixed) or manually prepared assay plates were utilized in another laboratory (Biological Research Centre, Szeged, Hungary; see **Supp. Text File 2** and **Fig. S5** for Semi-automated method). Single cell phenotypic analysis was performed with BIAS software as described^5^. The results are displayed in detail at **Supp. Text File 3** and **Fig. S8-S9**. Altogether, we processed 948 (583 FIMM) serum or plasma specimens collected in 2017 and 2020, with ethics approvals and informed consent (**Supp. Text File 2, Table S5)** from 890 (545 FIMM) donors, of which 181 (42 FIMM) were individuals with a positive SARS-CoV-2 RT-PCR and/or ELISA test result.

#### Assay *results*

As the results include thousands of data points, we developed automated scripts for the visualization of the quality assessment, as well as for the assay results. The visualization includes assay control data and plate-based heatmaps (**Fig. S4**). The results, allowing the observer to view images of each (patient) sample separately for all Ab/Ag scores, are visualized as interactive plots (plate-based; **Supp. Data 1**). The plots allow the clicking of the data point and opening of the image of the selected sample in a separate window for additional visual evaluation, and as heatmaps (**Supp. Data 2)**. These enable a fast digital (re)view of the results, independent of place and time, with quantitatively scored findings, and the fast prioritization of the serum samples for further tests, e.g., for a neutralizing assay or for the re-evaluation of technically challenging samples or borderline cases.

As shown in **Fig. 2a**, the assay could distinguish between serum samples from donors measured SARS-CoV-2 positive (COVID-19) and negative samples (Neg) in RT-PCR with a high level of significance as indicated by low p-values (**Table S6**). The *p*-values were lowest for the N and S proteins, for both IgA and IgG. IgM produced the least specific responses (**Fig. S6a, Table S6**), mostly explained by the appearance of unspecific staining in some samples. For IgA against the SARS-CoV-2 M protein, rarely employed in ELISA tests, our assay gave a high signal (i.e., positive ratio) for samples taken within <2 weeks of the positive RT-PCR test result (**Fig. 2a**). Based on these findings, the assay enables the simultaneous determination of different Ig class responses against multiple antigens. This is beneficial 1) for the separation of SARS-CoV-2 positive and negative cases, 2) for retrospective timing of the development of the infection and immunity against it, and 3) for distinguishing between vaccine- and infection-induced Ig responses.

**Figure 2.**
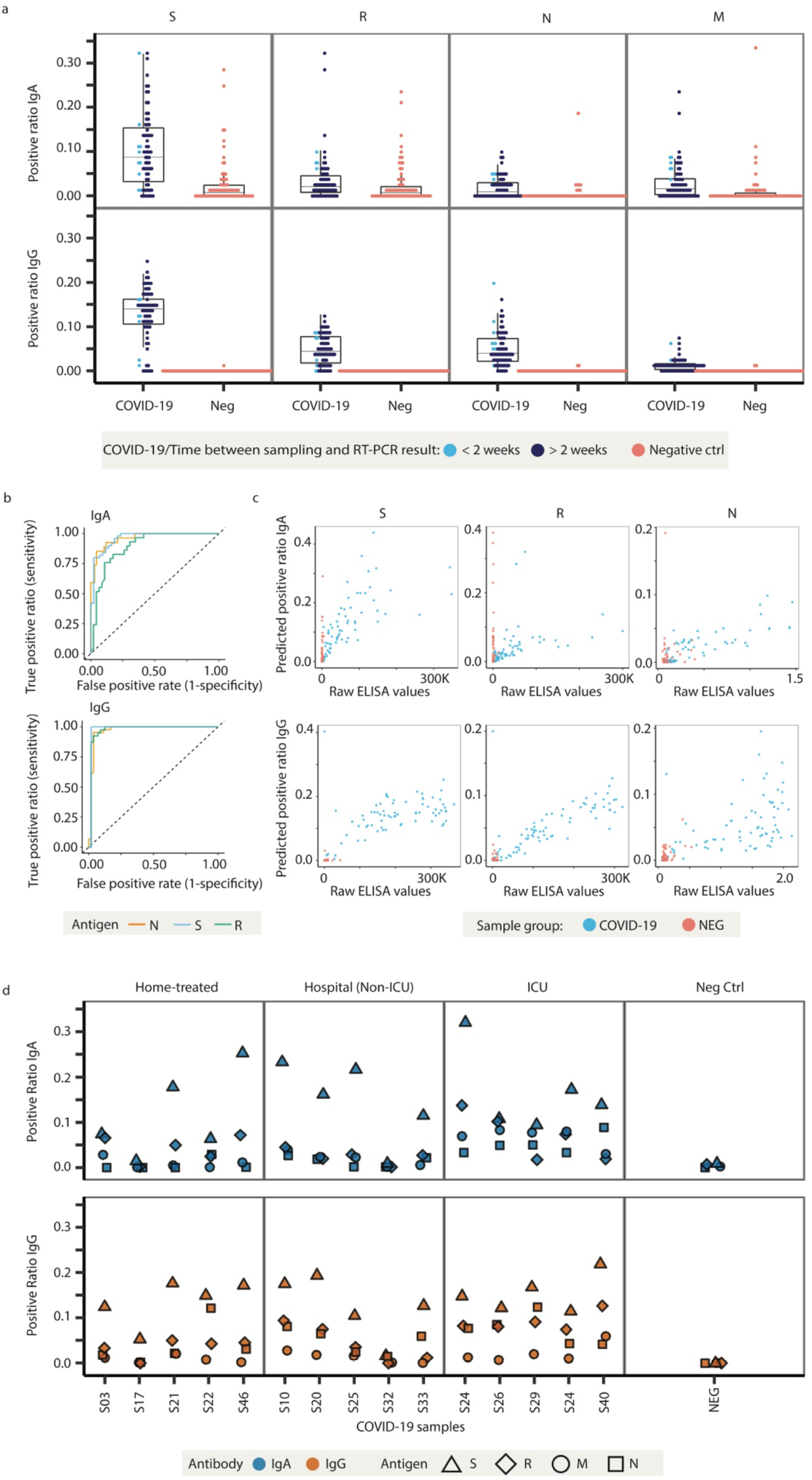
Performance of the assay. **a)** The dot plots show the distribution of sample prediction values of IgA and IgG antibody responses for S, R, N and M antigens present in the serum samples obtained from COVID-19 positive (COVID-19; collected in 2020) and negative (Neg; collected in 2017) patients. The sera obtained from COVID-19 positive patients are marked to show the time from a SARS-CoV-2 positive RT-PCR result. See Table S6 for the statistics. **b)** Comparison of the predicted values (IgG and IgA for N, S, and R antigens) with expert data, displayed as ROC curves. **c)** Correlation of the assay-predicted values (positivity ratios) with those of ELISA for IgG and IgA against the S and R antigens (high-throughput ELISA; n=42 COVID-19 patients, 80 samples; NEG n=80 patients, 80 samples), and against the N antigen (traditional ELISA; n=42 COVID-19 patients, 79 samples; NEG n=80 patients, 80 samples) antigens. ELISA values for the S/R vs. N antigen differ in scale, as they were obtained via chemiluminescence vs. colorimetric detection, respectively. See Table S7 for the statistics. **d)** Patient-specific antibody responses at different time points from the onset of symptoms or from obtaining a positive SARS-CoV-2 RT-PCR result plotted according to COVID-19 severity (treated at home, hospitalized/non-ICU, and hospitalized/ICU).

#### Comparison of expert opinion

An indirect immunofluorescence assay is commonly carried out on virus-infected cells fixed on glass slides, for which the findings rely on the expert’s visual inspection under a fluorescence microscope. To evaluate the quality of our per-well predictions, we performed a comparison against the consensus finding of six experts’ visual inspection. We randomly selected a balanced number of images (IgG/IgA x S/N/R plates) to represent the antibody reactivity for SARS-CoV-2 antigens of both expected positive (COVID-19 patient sera) and expected negative (samples collected in 2017, see **Table S5**) cases. Images from IgM and M-protein plates were not included due to unspecific or unclear staining patterns. Six experienced virologists/cell biologists annotated a set of 576 images as positive, negative, or unclear, one by one (**Fig. S7)**. 26 images for which the experts did not reach a consensus (majority vote: all IgA, with 23 opinions for IgA R-antigen) were removed from the evaluation. Model quality was assessed by measuring the area under the receiver operating characteristic curve (AUC) (**Fig. 2b**), which relates to the prediction’s sensitivity and specificity, and is considered optimal at a value of 1. For IgG, the model performed at an AUC of 0.98, 0.97, and 0.98, and for IgA it performed at an AUC of 0.96, 0.96, and 0.89 for S, N, and R antigens, respectively.

#### Comparison to ELISA assay results

Next, we compared our per-well predictions to the results of SARS-CoV-2 ELISA tests (IgG, IgA, and IgM against N, R and S, see **Supp. Text File 2** for method), performed as in^2,3,10^. The comparison used 83 samples from 42 patients (#F1a; #F1b) with confirmed SARS-CoV-2 infection detected by RT-PCR (**Table S5**) and 500 pre-COVID-19 samples collected in 2017 (#F2). Spearman correlations between the predicted positive ratio of our assay and the N, R, and S ELISA results were between 0.81–0.82 for IgG and 0.56–0.80 for IgA, while IgM demonstrated the lowest correlations characterized by values ranging between 0.17–0.44 (**Fig. 2c, Fig. S6b, Table S7, and Supp. Text File 2** for the Methods).

To evaluate whether severely ill patients would demonstrate a specific antibody pattern, we plotted the predictive positive ratios as a function of the severity of COVID-19 disease (treated at home, hospitalized/non-Intensive Care Unit-ICU, and hospitalized/ICU) (**Fig. 2d**). Interestingly, ICU patients had higher anti-N protein IgG and IgA levels as compared to the other groups, however, no definite conclusions can be drawn from these findings due to the low number of samples.

Altogether, our findings highlight the flexibility and the utility of the mini-IFA method, as well as the utility of the produced data.

## Conclusions

The automated mini-IFA method enables high-throughput screening of antibodies from a small volume of serum, with a performance comparable to ELISA, as exemplified here by the analysis of serum specimens from SARS-CoV-2-infected patients. The assay can simultaneously detect immune responses against multiple antigens and up to three Ig classes, thus it can potentially improve diagnostic accuracy compared to single-antigen tests. Antigen presentation by transient expression in cells allows 1) executing the assay in a laboratory of low biosafety level, and 2) the use of complex antigens, including those of any new virus variants, without the need of protein purification, thus it has the potential to overcome the limitations of conventional IFA (for 1) and ELISA methods (for 2). New ML model needs to be trained for each antigen. Here, we labelled on average 4,500 cells in each antigen-antibody combination which took approximately a workday from an expert. Cell -based format here was considered as the most physiological way to express the antigens, as well as to reveal the information on their cellular localization, which is not possible with ELISA or bead-based immunoassays. The benefit of the assay format over ELISA or similar methods is there being no need to produce and purify the protein as the cells are used to express the antigen. The additional benefits could include higher local antigen concentration, better folding (although potentially distorted by fixation), and detection of autoantibodies, as well as all the feature -based information the images can provide for scientific studies.

A similar approach has been earlier suggested^1^ and tested^11^ for SARS-CoV-2 using alive pathogen in BSL-3. Their analysis focuses on scoring the ratio of median antibody signal between infected and non-infected cells and using this ratio in ROC analysis to set an optimal threshold to deem sample either positive or negative for IgG, IgA, and IgM antibodies separately. However, in their set-up, the response to virus as such (Pape et al, 2020) does not reveal any specific responses to different virus antigens, whereas in our assay, the measurements on different virus antigens can be used to differentiate those only vaccinated (showing Igs only towards S protein/R antigen) and those who had the earlier disease (showing Igs also to other virus proteins).

Another benefit of our new methodology is the simplified visualization of the results which would enable a straightforward clinical application. End users of the assay can easily compare the visualizations of the quantitative results to sample images. This facilitates digital archiving, reduces bias caused by intra-/interobserver variability, as well as reduces microscopy workload/time, and increases the reproducibility of the results. Thus, the assay offers unique benefits compared to similar methods utilized for viral diagnostics, which use live viruses, manual immunostaining of slides, and visual inspection by an expert. The assay can be used, for example, for kinetic and longevity studies of antibody responses induced by infections, and it could serve as a useful post-vaccination tool for SARS-CoV-2 serosurveillance studies. Moreover, the assay could work with cells infected with entire viruses in the early stages of any pandemic before the relevant viral antigens are cloned. However, here we optimized the method for transfected antigens, as with those, in addition to alleviating users from the requirement of high biosafety levels, we can detect separate responses for each antigen, e.g. vaccinated from non-vaccinated but infected.

The method described here allows the simultaneous quantitative determination of responses against multiple antigens and different antibody classes in an automated fashion, something not achievable using the classical methods, such as ELISA, widely used in clinical practice. The combination of high throughput imaging and machine learning makes the proposed assay highly beneficial over existing serological assays because of its low cost, high sensitivity, and robustness. The richness of the data derived from the images of antibody-antigen interactions gives orders of magnitude more information, e.g., for studies of the immune responses to specific virus proteins, some of which cannot be expressed in any other system. This is beneficial for any vaccine development and understanding the immunogenic responses to infections.

As exemplified by the SARS-CoV-2, readily available, low-cost serology tests are key to control epidemics. Our proof-of-concept study for automated mini-IFA serodiagnostics, presented here, suggests that the assay translates to various pathogens with high-potential for wide-scale research and clinical applications.

## Supporting information

Supplementary measurement data

## Data Availability

All data produced in the present study are available upon reasonable request to the authors.

https://fimm-covid-19-hca.github.io/

http://86.50.253.10:8888/

https://dx.doi.org/10.5281/zenodo.6352550

## Data Availability Statement

Additional data are provided as supplemental data (tables, figures, and files). Scripts are available at the GitHub repository: https://github.com/fimm-covid-19-hca/mini-IFA_paper. Extracted single-cell features from microscopic images to train and test models are available at Zenodo: https://dx.doi.org/10.5281/zenodo.6352550

## Supplementary Information

The manuscript contains supplementary data: **a)** SI PDF file for Supplementary Text Files 1-3, Supplementary Figures S1-S9, Supplementary Tables S1-S8, as well as **b)** Supplementary Data 1 (https://fimm-covid-19-hca.github.io/; for description, see the the SI PDF) and **c)** Supplementary Data 2 (PDF file).

## Acknowledgements

The authors thank Minerva Institute (Helsinki, Finland) for providing utilities for the project; Prof. Perttu Hämäläinen (Aalto University, Finland) for providing expertise of his group for the project, the FIMM High Throughput Biomedicine Unit for providing access to high-throughput robotics, and the FIMM High Content Imaging and Analysis Unit for HC-imaging and analysis (HiLIFE, University of Helsinki and Biocenter Finland). We acknowledge support from the LENDULET-BIOMAG Grant (2018-342), from the European Regional Development Funds (GINOP-2.3.2-15-2016-00006, GINOP-2.3.2-15-2016-00026, GINOP-2.3.2-15-2016-00037), from the H2020-discovAIR (874656), from the H2020 ATTRACT-SpheroidPicker, and from the Chan Zuckerberg Initiative, Seed Networks for the HCA-DVP. The Finnish TEKES FiDiPro Fellow Grant 40294/13 (VP, OK, LP, PH), grants awarded by the Academy of Finland (iCOIN-336496: OK, VP, OV; 308613: JH; 321809: TS; 310552: LP; 337530: IJ, FIRI2020-337036: FIMM-HCA, AH, LP, VP, PH), the EU H2020 VEO project (OV) and Minerva Foundation for COVID-19 research project grant (VP) are also greatly acknowledged. CG is funded by the Academy of Finland Flagship programme, Finnish Center for Artificial Intelligence. OrthoSera Ltd. was funded by NKFIH grants (2020-1.1.6-JÖVŐ-2021-00010, and TKP2020-NKA-17). The authors thank Dora Bokor, PharmD, for proof-reading the manuscript.

## Contributions

JH, PH, and VP originally designed and led the project, and KrB, JH, PH, and VP supervised the project. JH, EM, VP, MP, and LT designed the laboratory experiments and JH, MH, AK, MP, EGyS, and LT executed the experiments. IJ and LKa provided unpublished constructs. KaB, AKa, LKe, GT, AV, and OV provided serum samples; EF and ZsL collected 165 positive sera with patient data and performed ELISA analyses in Hungary in co-operation with Orthosera Ltd. AH, EM, LP, and MP performed high-content imaging and JH, IJ, PK, LL, ES, and TS analysed IFA samples. CG, FK, and LP developed the analysis pipeline. CG and LP facilitated the prediction model selection and its comparison to ELISA and expert annotations. PH, AK, and FK developed the image analysis software (BIAS). RH prepared the model for nuclei detection and segmentation. SP prepared the QC and visualization scripts. AD, EM, and MP performed expert training. CG, JH, MH, PH, LP, MP, and VP wrote the main parts of the manuscript. AD, CG, MH, LP, MP, SP, VP and LT prepared the figures, tables and data files included in the paper. All authors read, reviewed, and approved the final version of the manuscript.

