## Supplementary methods and data for "Image-based & machine learning-guided multiplexed serology test for SARS-CoV-2"

**This PDF file includes:**

Supplementary Text Files 1-3 (SI References are included directly to the text files)
Supplementary Figures S1 to S9
Supplementary Tables S1 to S8

**Other supplementary materials for this manuscript include the following:**

Supplementary Data 1 shows the visualization of quantitative sample-specific IgG results and their distribution compared to controls and other samples, together with microscopic images. It is located at https://fimm-covid-19-hca.github.io/ . The webpage contains separate files (.html) for interactive detection of samples' IgG responses towards SARS-CoV-2 antigens (S, R, N, M) on 384-well plates (S01-S04) with corresponding microscopy images. The graphs show the positive ratio (Y-axis on the left; IgG) for IgG with S/R/N/M antigen for each patient sample (X-axis; Sample names). Regions of the graph can be zoomed in, and the individual data points can be hovered to view 1) the microscopic image for a IgG/Ag pair for each sample and 2) the positive ratios of the antigen with different Igs (IgA, IgG, IgM) for each sample. The results shown are created with the automated mini-IFA assay, and include donors' serum sample sets #F1a, #F1b, and #F2 (see Table S5). Images of controls utilized in analysis pipeline are also shown. (https://fimm-covid-19-hca.github.io/)

Supplementary Data 2 (PDF file, separate)
The heatmap of results implicates the positive cell ratios for different IgA/IgG & antigen M/N/R/S pairs in each serum sample.

**Supplementary Text Files 1-3**

**Supplementary Text File 1. Protocol for Pietiäinen et al. Image-based & machine learning-guided multiplexed serology test for SARS-CoV-2**

1. ***Sample preparation***

*1.1* ***Serum transfer to ECHO Source plates***

1.1.1 Plan Sample Src plates so that there's room for controls, or use separate plate for control samples.

1.1.2 Thaw 96 well plates (polypropene plates if possible) with serum from -20 degrees, centrifuge 10 min 700 g.

1.1.3 Transfer with automated pipettor 50 µl volume to Labcyte 384PP plate.
- 4 x 96 well plates to 1x 384
- 96 tips, new box for each serum plate

1.1.4 Transfer Hoechst 250 nl/well with acoustic or other sub-microliter dispenser
(Echo 525 buffer calibration *384PP_AQ_BP*)

1.1.5 Seal plates, shake for > 30 mins high speed.
Put it back in the freezer or continue with the IFA assay.

2. ***Transfection of VERO E6 cells***

**2.1. VERO E6 cell culture**

2.1.1. Culture cells in T-75 cell culture flasks until they reach 90-95% confluency (cells from one flask are enough for transfection of 3-4 384-well plates)

2.1.2. Aspirate the cell culture medium and rinse the cells with 3 ml of Trypsin-EDTA solution

2.1.3. Aspirate the Trypsin-EDTA solution and add 2 ml of Trypsin-EDTA to detach the cells

2.1.4. Incubate for 2-5 min in 5% $CO_2$ +37°C incubator, until cells detach

2.1.5. Resuspend the cells from one flask to 38 ml of cell culture medium

2.1.6. Calculate the cell density of the cell suspension and adjust if needed (the density should preferably be 1.2-1.6 x $10^5$ cells/ml = 3000-4000 cells/well)

Note: If more than one flask of cells is needed, trypsinize all the cells and mix them together to generate a uniform cell suspension for the transfections. Keep the cell suspension on rotation to prevent cell aggregation.

*2.1* **Transfection**

2.1.1 Calculate the volume of OptiMEM solution, plasmid DNA (S-pCAGG, RBD-pCAGG, N-pCAGG and M-pEBB), Fugene HD and Vero E6 cell suspension needed for each transfection according to the following formulas (n = the number of plates to be seeded/construct):
500 µl of OptiMEM x (n + 0.5)
5.3 µg of plasmid DNA x (n + 0.5)
18 µl of Fugene HD x (n + 0.5)
10 ml of Vero E6 cells x (n + 0.5)

2.1.2 Pipet the calculated volume of OptiMEM into a 15 ml conical tube (separately for each construct)

2.1.3 Add the calculated volume of plasmid DNA into the OptiMEM solution and mix well

2.1.4 Add the calculated volume of Fugene HD and mix well by pipetting up and down

2.1.5 Just before cell seeding, mix the needed amount of cell suspension with the transfection mix in a separate tube/bottle and incubate at RT for 15-30 min
Note: Keep the transfected cells on rotation to prevent cell aggregation.

2.2 **Cell seeding**

2.2.1 Prepare an automated dispenser for cell dispensing. Wash with water, 70% EtOH, water, PBS before and after dispensing

2.2.2 After washing prime with medium and before each construct seeding prime with cell suspension

2.2.3 Dispense 25 µl to all wells of sterile 384 well plate with lid suitable for imaging

2.2.4 Incubate cells in the cell incubator for 48 h at +37° C with 5 % $CO_2$

***3 <u>Fixing the cells</u>***

**3.1 8% PFA is added with an automated dispenser. PBS washes are performed with a plate washer dispenser.**

***3.1.1 Make sure 8% PFA is free of precipitate before dispensing***

3.1.2 Prepare automated dispenser and dispense 25 µl of 8% PFA on top of cells (Collect toxic PFA waste)

3.1.3 Incubate 10 min RT

3.1.4 Wash the plate twice with 50 µl PBS and leave 70 µl PBS for storage.

3.1.5 Wrap the plates to parafilm and store at +4 °C.

**4 *<u>Permeabilization, blocking and staining</u>***

**4.1 Prepare Acoustic or other sub-nanoliter transfer protocols and liquids in advance**

4.1.1 Add controls to serum sample plates, add them to transfer lists (800 nl for 1:25 dilution and 200 nl for 1:100 dilution)

4.1.2 Controls to serum plates (384PP): spin serum samples extensively to precipitate any debris. Take the supernatant volume needed and add Hoechst 1/200. Each sample is needed 1 µl/ well + 20 µl. Spin plates briefly when done

4.1.3 Prepare buffers and other liquids needed for the number of plates run:

4.1.3.1 **Wash buffer**: TBS 165 ml/plate + dead volume (instrument w)

4.1.3.2 **Blocking and permeabilization buffer**: 3% BSA-TBSTx (0.25%) 15.4 ml /plate + dead volume (instrument d1)

4.1.3.3 **Assay buffer**: 0.5% BSA-TBS 6.4 ml /plate + dead volume (d2)

4.1.3.4 **Backfill buffer**: Hoechst (1/200) in assay buffer for backfill 122 µl /plate + dead volume (instrument d3)

4.1.3.5 **Secondary Abs in assay buffer**: 5.5 ml /plate + dead volume (instrument d4)

4.2 **Blocking and permeabilization buffer is added with automated dispenser (d1) and washes performed with plate washer dispenser (w), then buffer for serum samples (d2) and backfill (d3) are performed with automated dispenser**

4.2.1 Prepare instruments for appropriate liquids (d1, d2, d3, w)

4.2.2 Washer dispenser aspirates plate empty (5 µl left behind) and adds 40 µl Blocking and permeabilization buffer (w)

4.2.3 Incubation 15 min at room temperature
4.2.4 Washer dispenser (w) washes plate 2 times with 50 µl of Wash buffer and aspirates plate empty (5 µl left behind)
4.2.5 Automated dispenser (d2) dispenses 15 µl Assay buffer to all wells
4.2.6 Automated dispenser (d3) dispenses Backfill buffer to serum dilution wells so that each well contains same amount of Hoechst in the end (dispensing volumes 0/600/800 nl)

4.3 **Serum samples are added with Acoustic dispenser or similar**
4.3.1 When plate is ready from previous steps spin the plates briefly
4.3.2 Dispense serum samples to assay plates, 800 nl for 1:25 dilution and 200 nl for 1:100 dilution
4.3.3 Incubation for 1.5 hours at room temperature

4.4 **Secondary antibody is dispensed with automated dispenser (d4), washer dispenser (w) is washing the plate**
4.4.1 Prepare the automated dispenser (d4) with secondary antibody solution, washer dispenser (w) should still have buffer in it
4.4.2 Washer dispenser washes plates three times with 50 µl TBS (collect toxic Hoechst waste)
4.4.3 Dispense 15 µl of secondary antibody solution to each well
4.4.4 Incubate 45 min at room temperature
4.4.5 Wash with washer dispenser (w) two times 50 µl TBS, leave with 70 µl of TBS

5 ***Plates are ready for imaging, or can be stored at +4C wrapped in parafilm***

6 ***Imaging***
6.1 Imaging is performed in RT with PerkinElmer Opera Phenix high-throughput spinning disc confocal microscope using Harmony 4.9 software and two Andor Xyla sCMOS cameras

6.2 Basic settings
6.2.1 Plate setting: PerkinElmer 384 CellCarrier Ultra flat bottom plate
6.2.2 Two-peak autofocus in confocal mode with binning 1
6.2.3 20x water-immersion objective (NA 1.0)

6.3 Channel settings
6.3.1 405 nm laser (Hoechst), 435-480 nm emission filter, 100 ms exposure time, 100 % laser power
6.3.2 488 nm laser (IgM), 500-550 nm emission filter, 100 ms exposure time, 100 % laser power
6.3.3 561 nm laser (IgG), 570-630 nm emission filter, 200 ms exposure time, 100 % laser power
6.3.4 640 nm laser (IgA), 650-760 nm emission filter, 200 ms exposure time, 100 % laser power
6.3.5 405 nm channel is separated from both 488 nm and 647 nm in order to scan 405 nm and 568 nm on camera 1 as well as 488 nm and 640 nm on camera 2 to minimize bleed through between the channels

6.4 Layout selection
6.4.1 A grid of nine adjacent fields selected from the center of the well, 5 % overlap between the images. Area of the selection is 3.83 um2 or 36 %

of the total well area. Image size is 2160 x 2160 pixels with 6.5 x 6.5 um pixel size

6.4.2 Z-stack of 2 fields selected, first plane at -4.0 µm, 3 µm distance between planes

6.4.3 Imaging time is 2 h 6 min per one full plate

6.5 Image export

6.5.1 Measurements are exported with the “Export data” -function in data management settings. Choose option “Measurements” to export raw tiffs and the index.idx metadata file for BIAS import.

### 7 ***<u>Image analysis and quality control in BIAS</u>***

7.1 Image pre-processing

As quality control well-focused images are created and uneven illumination is corrected.

7.1.1 Focused images are retrieved by maximum intensity projection on z-stack images acquired in multiple focal planes with the “Flatten” module using the “MIP” method.

7.1.2 Flat-field correction is applied to fix non-uniform illumination with the CIDRE algorithm using the “Illumination correction” module: additive noise was eliminated and acquired intensity gain is compensated for.

7.2 Segmentation

7.2.1 Single nucleus objects are segmented with a Mask R-CNN deep convolutional neural network model (trained with the nucleAIzer method) on the Hoechst channel in the “Segmentation” module using the “Deep neural network” option and the “Generic nucleus segmentation” model.

7.2.2 Cell regions are determined in the “Mask operators” module as 7 µm extensions (dilation) of the nucleus objects starting from the given nucleus contour each as far within the 7 µm radius as such adjacent extensions would not overlap.

7.2.3 For each cell, its enclosed nucleus segmentation is subtracted from its region to yield a cytoplasm area, also in the “Mask operators” module using the “Complement” option.

7.3 Feature extraction

Single-cell level features were measured in the “Feature extraction” module by each segmented object (nucleus, cell, cytoplasm).

7.3.1 Morphology is measured on the segmented regions as size and shape descriptors such as area, perimeter, eccentricity etc.

7.3.2 Intensity features are calculated from the intensity image corresponding to the given channel, considering only the region covered by the given object, for each channel and object, respectively. Basic statistics (e.g. mean, median, minimum, maximum, sum) of intensities are measured.

7.3.3 Measured texture features correspond to the so-called Haralick-features that describe the pattern properties of a region corresponding to a segmented object, for each channel and object, respectively, similarly to intensity descriptors.

### 8 **Train models and predict new data**

8.1 Detailed description of how to train models and predict new data is available in a Jupyter Notebook, see https://github.com/fimm-covid-19-hca/mini-IFA_paper; code share at GitHub.

9 **Quality control and visualization**

9.1 R scripts used for QC and visualization are shared in GitHub repository, see https://github.com/fimm-covid-19-hca/mini-IFA_paper.

**Supplementary Text File 2.** Supplementary Materials and Methods.

### Supplementary Materials and Methods

The detailed info of all materials and reagents used is given in **Table S1**. The automated assay pipeline protocol is described in **Supp. Text File 1**.

#### Cell line, and plasmids

The study made use of the following plasmids for recombinant expression of three SARS-CoV-2 proteins and receptor binding domain (RBD; or R) of the spike (S) protein: S protein with His-tag (S-pCAGGS; described in[1]), RBD with His-tag (RBD-pCAGGS; described in[1]), membrane (M) protein with HA-tag (M-pEBB; HA-Tag) and nucleoprotein (NP; or N) with His-tag (NP-pCAGGS; described in[2]) (see also Supp. Table S1). For M-pEBB, M gene, according to hCoV-19/Finland/1/2020 (GenBank accession MT020781) sequence, was synthesized and cloned into pEBB-N-HA mammalian expression vector by GeneArt (Thermo Fisher Scientific).
Vero E6, African green monkey kidney cell line (ATCC; mycoplasma tested), were grown in Minimal Essential Medium Eagle (SIGMA, US) supplemented with 10% fetal bovine serum (Gibco, US), 2 mM L-glutamine (Gibco), 100 IU/mL penicillin and 100 µg/mL streptomycin (Gibco/Sigma). Vero E6 cells were cultured to a 90-95% confluence in T-75 cell culture flasks and detached using 0.25% Trypsin-EDTA (Gibco).

#### Patient serum samples and ethical permissions

Information of patient samples is provided in **Table S5**. For automated IFA-assay (Helsinki, Finland), the SARS-CoV-2 patient sample panels 1&2 comprised of 83 blood samples (serum, plasma) drawn from 45 individuals, of whom 42 had been tested positive in clinically validated SARS-CoV-2 RT-PCR test (HUSLAB, Helsinki University Hospital Laboratory Diagnostics); the panel also included three samples from COVID-19 suspects with negative/no result in SARS-CoV-2 RT-PCR. The RT-PCR testing was performed from nasopharyngeal swab samples. The serum samples from COVID-19 patients were drawn 8 to 81 days after onset of symptoms. The data and samples for this panel were collected under research permit HUS/211/2020 and ethics committee approval HUS/853/2020 and HUS/1238/2020 (Helsinki University Hospital, Finland). A panel of 500 serum samples collected during 2017 from patients with suspected Puumala virus infection served as SARS-CoV-2 antibody negative controls (Research permits HUS/167/2016, HUS/38/2018 and HUS/244/2021). Written informed consent was obtained from all participants. The study was conducted in accordance with the Declaration of Helsinki.

For semi-automated mini-IFA assay, a total of 200 negative control sera and 165 positive samples derived from 145 patients were provided by the Hungarian National Blood Transfusion Service, the Department of Dermatology and Allergology of the University of Szeged and the Orthosera Ltd. Samples predicted positive were collected from 139 RT-PCR-and/or ELISA-confirmed patients and from 6 further patients having symptoms or tested by another method under general informed consent. Sera were stored at -80°C and heat inactivated at 56°C for 60 min before the assay.

Sample collection and this study was approved by the Scientific and Research Ethics Committee of the Hungarian Medical Research Council (clearance numbers IV/3457-2/2020/EKU and IV/3757-4/2020/EKU). The study was conducted in accordance with the Declaration of Helsinki.

**Automated mini-IFA assay**

The assay pipeline protocol is provided in **Supp. Text File 1.**

**Transfection.** Vero E6 cells were transfected using FuGENE HD (Promega, US) at 3.5:1 reagent to plasmid ratio. The FuGENE HD-plasmid mixes were prepared in Opti-MEM (Gibco) following the manufacturer's protocol, after 10-15 min complex formation, a suspension of trypsinized Vero E6 cells in fully supplemented media was added, the resulting suspension was incubated at RT for 15-30 min in rotation, and the cells were seeded onto plates either manually or with automation.

In the automated assay, the transfected cells were seeded to CellCarrier Ultra 384-well microplates (PerkinElmer, US) with a seeding density of approximately 3-4 x 10^3 cells/well in 25 µl per well using MultiFlo FX RAD dispenser (BioTek, US) and incubated at 37°C humidified atmosphere for 48 h in the growth media. Cells were fixed in 4% PFA for 15 min at RT, washed twice with PBS, and filled with fresh PBS. The plates were stored at 4°C prior to use. Liquid dispensing and aspiration were performed using EL406 -plate washer/dispenser (Biotek).

**Immunostaining.** Washing, liquid dispensing and aspiration were performed with EL406, buffer dispensing with Certus FLEX 0.3 (Gyger) and serum dispensing with Echo 525 (LabCyte). Cell permeabilization and blocking of the unspecific binding were performed using 0.25% Triton X-100 in Tris-buffered saline (TBS) (0.05 M Tris-HCl, 0.15M NaCl, pH 7.6), containing 3% bovine serum albumin (Biowest, US) for 15 min at RT. Cells were washed three times with TBS before serum samples were applied on wells. Two different dilutions of each serum sample were used; 1:25 (for detection of IgA; IgM) and 1:100 (for detection of IgG) in 0.5% BSA in TBS with 1:5000 dilution of Hoechst 33342 (Lifetech) and incubated at RT for 1.5 h. After washing steps, the fluorochrome-conjugated secondary antibodies, goat anti-human IgG DyLight 550 (1:500; Invitrogen, US), goat anti-human IgM AlexaFluor (AF) 488 (1:1000, Invitrogen) and goat anti-human Serum IgA AF647 (1:1000; Jackson ImmunoResearch, US), were used for detecting different immunoglobulins from patient sera by incubating them in the wells for 45 min at RT. To determine the transfection percentage, mouse anti-His-tag antibody AF647 (1:1000, BioLegend, US; for S, N, and R-protein transfections), mouse anti-HA tag AF488 (1:1000, Invitrogen; for M-protein transfection) and in-house rabbit anti-N and anti-S/R stained with AF488 secondary antibody (1:1000, Invitrogen) were used. All assay plates had five wells reserved for the transfection controls. For more accurate calculations of transfection efficacy, a parallel set of plates with transfected Vero E6 cells were stained with antibodies for each antigen as described above, and a transfection control and mean transfection percentages were calculated from images captured as described below. The transfection percentage was determined by selecting a mean intensity cut-off for immuno-stained transfected cells and calculating the percentage of transfected cells of all cells. The transfection rates for N, R, S, and M are shown in **Table S2**.

**High-content imaging.** Opera Phenix confocal spinning-disk high content screening microscope (PerkinElmer, Inc., Waltham, MA, USA) was used for imaging of 384-well plates for automated IFA-assay (High Content Imaging and Analysis Unit, FIMM, HiLIFE, University of Helsinki, Finland). Screening was conducted with a 20x water immersion objective (NA 1.0, working distance 1.7 mm, depth of focus 1.8 µm, effective xy resolution 0.66 µm) and four excitation lasers (405 nm with emission band-pass filter 435/480; ex 488, em 500/550; ex 561, em 570/630; ex 647, em 650/760).

Nine fields-of-view with 5% overlap were imaged per well using two predetermined Z focus planes with laser-based autofocusing. The images were captured with two Andor Zyla sCMOS cameras (16-bit, 2160 x 2160 px, 6.5 µm pixel size).

**Semi-automated mini-IFA assay**

First, we tested the protocol in 96-well plate set-up for the first 40 samples, with manually transfected plates (for transfection efficiency, see **Table S2**). In the manual protocol, after 15 min incubation with the transfection mix, the cells were seeded to black 96-well tissue culture plates (Greiner, Austria) with a density of 12,000 cells/well. After 48 h incubation, culture medium was removed and the cells were washed with DPBS, fixed with 4% PFA for 10 min at RT, washed again with DPBS twice and stored in DPBS at 4°C. For the manual protocol, 8- and 12-channel pipettes and 50 ml reagent reservoirs were used.

Additional 325 samples were investigated on ready-to-go plates produced by the automated method. Permeabilization was performed in 3% BSA with 0.2% Triton X-100 in TBS for 5 min at RT and blocking was done in 5% BSA in TBS for 60 min at RT. Serum samples diluted in 1:25 ratio in 0.5% BSA in TBS were incubated on the plates for 90 min at RT. AF546-conjugated anti-human IgG (1:500; Invitrogen) along with AF488-conjugated anti-human IgM (1:1000; Invitrogen) or AF647-conjugated anti-human IgA (1:1000; Jackson ImmunoResearch) were applied for 120 min at RT. The 6x-His Tag mouse antibody (1:1000; Invitrogen) was used for overnight at 4°C and AFPlus488-conjugated anti-mouse IgG (1:500; Invitrogen) or AFPlus647-conjugated anti-mouse IgG (1:500; Invitrogen) was added for 60 min at RT to detect the His-tagged virus antigens (S, N, R) in the transfected cells in each well. Each antibody was diluted in 1.2% BSA in TBS. Nuclei were counterstained with 200 ng/ml DAPI for 15 min at RT. Wells were washed with TBS 3 times for 5 min between each step and covered with TBS for imaging.

Images were obtained using the Operetta high-content imaging system (PerkinElmer) with a 20x air objective (20 × long WD; NA 0.45, working distance: 7.8 mm; depth of focus: 4.6 µm). Images were acquired with a Peltier cooled CCD camera (14-bit, field of view: 675 × 509 $\mu m^2$; optical xy resolution: 0.67 µm). Six fields of view were acquired per well using one Z focus plane determined by the laser-based autofocus system. For detecting fluorophores, the following excitation and emission filters were used: DAPI (380/40; 445/35); Alexa Fluor 488 (475/30; 525/50); Alexa Fluor 546 (535/30; 595/70); Alexa Fluor 647 (630/20; 675/50).

**Image processing and feature extraction**

Images obtained by both methods described were processed using BIAS software[3,4]. Pipeline created for the analysis consisted of three main steps: 1) pre-processing of the images, 2) segmentation and 3) feature extraction. In the pre-processing, a maximum intensity projection was created from each stack of images in different focus depths. Non-uniform illumination was corrected separately for each channel using the CIDRE method[5]. Deep learning segmentation method[6] was applied to segment nuclei in images. From these nuclei regions, two additional segmented regions were defined: 1) the cells were defined by dilating nuclei regions with maximum 7 µm radius so that adjacent cells did not overlap, and 2) cytoplasm regions were defined by subtracting nuclei segmentation from the cell segmentation. Finally, morphological properties of these three different regions as well as intensity and texture features from all channels were extracted (in total 255 features) for cell classification.

**Single-cell phenotyping annotations (classes, numbers, distribution in cells)**

For the automated method, we employed supervised machine learning to predict five different cell types: Positive (P), Negative (N), Atypical (A) and Small Bright (S) cells as well as other artefacts that can be considered Trash (T). These classes were chosen based on clinical diagnostics classification of microscopic findings in virological IFA assay (P, N, A, T) as well as to exclude e.g., rounded or dying cells (S). Cells with evenly distributed high specific intensity across the whole cells were labelled as Positive, whilst cells with low intensity were labelled as Negative. Cells with abnormally small, bright nuclei and a strong intensity of unspecific antibody staining were considered as Small Bright, to differentiate the dying or dividing cells from the true positive cells. Cells with atypical staining patterns, for example strong nucleus intensity, were considered as Atypical. We include an “atypical” class to capture cells that phenotypically resemble positives, but present unusual staining patterns compared to the positive controls, e.g. strong nucleus intensity. Moreover, atypical cells show higher non-specific fluorescence, and their classification thus serves as additional quality control to highlight unspecific antibody binding. Small, segmented areas, caused by the background of the serum samples were labelled as Trash.

To gather training data for our predictive model, the cells were annotated using an active learning feature within the BIAS software. We thus employed a simple machine learning setup to generate data for training a more sophisticated predictive model later on. For active learning, a Support Vector Machine (SVM) classifier within BIAS was trained on a set of initial annotations to classify samples from outside the training pool. The annotator then iteratively chose candidates from these new samples to extend the training data. The annotator then retrained the active learning model, allowing for a gradual increase in classification quality. Additional samples were added to the training set until a cross-validation accuracy of at least 0.85 per antigen and antibody was achieved. The annotations were initially performed by one expert to ensure consistency, and then checked against by other experts. We instructed the annotators to aim for a similar number of annotations in each of the five categories. Yet, the negative class includes approximately double the number of annotations than other classes, as it is the most typical class in the data. The annotators tried to focus on increasing the number of rare cases (atypical, small bright, trash) to make the training dataset more balanced.

In total, we annotated 55 496 cells across four plates from two experimental batches, covering four viral antigens and three antibody classes (**Table S3**). Roughly the same number of cells were annotated per antigen. The number of annotations vary slightly between plates, as sometimes more annotations were required to surpass the 0.85 cross-validation accuracy threshold. They vary more strongly between cell classes due to the frequency of specific examples in the respective samples.

**Feature normalization.** We included an additional quality control for feature normalization based on control wells as not all control samples were transferred correctly into plates. As the serum sample and nuclear stain (Hoechst) were transferred simultaneously, we based the quality control on the mean Hoechst signal in nuclei. We measured the mean and the standard deviation of Hoechst intensity from nuclei mean intensities in each well of a plate and ruled out all control wells that were not inside [mean - 1.0*std, mean + 1.0*std] range. The range was defined by optimizing the multiplier of std using wells that were known not to have correctly transferred samples. We found that mean +- 1.3*std was the most optimal range but to make the rule stricter we decided to use 1.0 as a multiplier.

Based on the controls, we normalized the image features per-plate to allow for model training and prediction across plates with potentially varying signal strength. As a common requirement for some of our machine learning models, we moreover standardized the features to approximately zero mean and unit variance. We compared three techniques, each drawing on image features from different sets of control samples:

- PosNeg: All transferred positive and negative controls.

- PosNegBal: The same amount of positive and negative transferred controls, based on sampling n controls from the set of transferred positive and negative controls respectively, with n being the highest number of transferred controls in both sets.
- NegOnly: Only negative transferred controls.

(PosNeg) allows us to obtain a data sample which is as representative as possible for the signal in positive and negative samples, respectively. However, the number of successfully transferred wells in either set might vary, which can lead to a shift of the mean towards a lower or higher signal. We hence evaluated (PosNegBal) where both types of controls contribute equally to normalization and standardization. As the signal in negative controls present mostly the background variation of the assay whereas the positive controls have more variation due to biologically meaningful differences, and because obtaining positive control samples in the first place can be more difficult, we also evaluated (NegOnly) which only leverages the set of negative controls. The best approach was determined based on its performance in per-cell predictions, assessed through cross-validation.

**Training, models, and used hyper-parameters (single-cell predictions).** We trained and evaluated three different types of machine learning models commonly used for the multi-class cell phenotype classification, drawing on the Python implementation in Scikit-learn[7]. For each classifier, we performed a comprehensive parameter grid search to determine the best model. Following an initial, coarse search, we fine-tuned the following model-hyperparameter configurations:

- Random Forest (RF). We search the number of decision trees, $\mathrm{N} \in \{50,100,200,300,500\}$, to avoid an overfitting of the estimator while keeping the estimator complexity low. We moreover consider different numbers of features for each tree node $\mathrm{M} \in \{\#\mathrm{features}, \sqrt{\#\mathrm{features}}\}$to trade-off the reduction of variance with the increase of bias.
- Support Vector Machine (SVM) with a radial basis function kernel. We evaluated the regularization parameter $\mathrm{C} \in \{0.1,1,10,50,100\}$ to achieve a good trade-off between a smooth decision surface, hence avoiding overfitting, and the correct classification of all training samples. We moreover evaluated different values of the kernel parameter $\gamma \in \{1.0, 1 \times 10^{\mathrm{n}}, 5 \times 10^{\mathrm{n}} \mid \mathrm{n} = -1, \dots, -4\}$ which controls the influence of individual training examples and thus the complexity of the decision boundary.
- Multilayer Perceptron (ANN) with logistic activation functions. We compared neural network architectures with one to three hidden layers of varying size$\{(256,128,64), (128,64,32), (256,128), (128,64), (64,32), (32,32), (128), (64), (32)\}$ controlling the model's complexity. We moreover evaluated regularization parameters $\alpha \in \{1.0, 0.1, 0.05, 0.01, 0.005, 0.001\}$to avoid overfitting.

We avoid model bias through class imbalances by including class weights in the training. For each antigen-antibody combination, we selected the best model based on a cross-validation of cell type predictions (**Table S4**).

**Cross-validation.** For the practical use of this assay, it is essential that the trained model can produce predictions on new, previously unseen plates for which no training data is available. To evaluate the prediction quality of different model candidates for use in per-well predictions, we consequently performed a four-fold leave-one-out cross-validation on the level of plates. It has also been used to identify the best normalization technique for cross-plate prediction.

Each cross-validation fold comprised training data of three plates and used the remaining plate as validation data. The ratio of training to validation data per fold was thus 3:1 (e.g., for S/IgG 3920:1301; **Table S3**). We calculated the sensitivity, specificity, and accuracy of predictions, and selected the best performing model (and normalization approach) based on the mean

$\text{sensitivity} \times \text{specificity}$ across all folds. This implements a maximum ignorance assumption on the later clinical use. If an application would benefit to trade-off one factor against the other, the model can be re-trained based on a corresponding weighting of both measures. The cross-validation was performed individually for each antigen-antibody combination, and the results for all tested classifiers and normalization approaches with optimized hyperparameters (**Table S8**) are presented in **Table S4**. We also show the confusion matrices for the ANN classifier for NegOnly and PosNegBal normalization scheme in **Fig. S3a and S3b**.

**Visual Inspection Data.** Following the single cell cross-validation analysis we proceeded to evaluate how well the selected optimal model works in the sample positivity prediction task. In this task we wanted to evaluate 1) how well the predicted positive cell ratio correlates with the ground-truth positivity measurement of the image data prepared with the presented workflow, and 2) the performance of the model on the positivity prediction task using the same images as ground-truth.

We hence performed a separate labelling study to obtain a second type of ground-truth data, reflecting expert virologists' judgements of sample positivity and negativity based on visual inspection of the very same images that are used for predictions in the proposed assay. The study was designed to closely mimic standard clinical practice. Six experts in immunofluorescence microscopy were asked to rate each in a selection of samples, shown next to a positive and negative control, as either 'positive', 'negative' or 'unclear'.

The sample images were procedurally selected and processed to remove experimenter bias (**Fig. S7**). For each combination of the IgG and IgA antibodies and the N, S, R antigens, we randomly picked 96 sample images from the test dataset. We selected the same amount of 24 samples from each of the four plates in the set. We moreover enforced a distribution of 50% assumed (samples taken before the virus outbreak vs. RT-PCR confirmed COVID-19 patients) positive and negative samples in each category to allow for well-differentiated sensitivity / specificity curves in the later evaluation. Each image was transformed using the same color transfer function, and only included the Hoechst 33342 channel for nuclear stain and one antibody channel. The positive and negative controls were specific to a plate and antibody/antigen combination.

The study materials were also procedurally composed, which allowed for straight-forward randomization of samples per participant to account for ordering effects. A screen calibration image was included in the beginning of the study material pack to ensure that all participants experience the sample and control images similarly. Each participant rated the same set of 576 samples, and the results were parsed automatically later to avoid any errors in translating the findings. Participation in the study took approximately 2 hours per participant. No sample was overlooked, resulting in a total of 1884 'negative', 1412 'positive', and 160 'unclear' ratings.

We assessed the inter-rater reliability, i.e. the degree to which our participants agreed with each other about their ratings, based on Fleiss' Kappa, an extension of Cohen's Kappa to more than two raters[8,9]. Calculated on ratings for the three classes 'negative', 'positive' and 'unclear', we found kappa = 0.71, i.e., very good[10] reliability. Following this analysis, we removed samples that were rated 'unclear' at least as often as they were rated 'positive' or 'negative'. This only applied to 26 samples, i.e., the final dataset contains 550 samples. We calculated the mean score for each sample (between 0=negative, 1=positive), ignoring 'unclear' ratings. Only 16 samples have a mean rating between 0.4 and 0.6, indicating a good consensus of sample positivity and negativity throughout. We finally binarized the results by considering any sample with mean score > 0.5 as positive.

**Per-well predictions.** We used the best cell type classifier and normalization scheme determined in the cross-validation to generate binary predictions of antibody reactivity for a specific antigen and antibody. To this end, we applied the selected model to all segmented cells identified in the respective sample well. Our per-well predictions are given as the positive ratio as the number of positively classified cells divided by the total segmented cell count in the well.

#### Comparison to Visual Inspection Data

We compared our predictions to the visual inspection data obtained from human experts as ground truth for this assay. We expect different assay applications to favor different trade-offs between assay sensitivity and specificity and hence do not report a fixed value. Instead, we highlight all possible trade-offs between these metrics by plotting the receiver operating characteristic (ROC) curve. Each point on the curve reflects a different threshold to binarize our predicted positive ratio for comparison with the binarized visual inspection ground truth. This threshold is a model hyperparameter and has an intuitive interpretation: it defines how many cells in the well image must be predicted positive to deem the entire well positive. For application of the assay, the threshold can be chosen arbitrarily or by evaluation on a set of validation ground truth data to obtain a specific sensitivity-specificity trade-off. We also report the area under this curve (AUC) as a scalar, aggregate measure of classification performance across all possible thresholds (see **Fig. 2b, Fig. S7**).

#### Comparison to ELISA

We compared our per-well predictions with ELISA results for the N, R, and S antigens of a sample subset across several test plates. ELISA represents an alternative to our assay, but, in contrast to our visual inspection data, cannot be considered ground truth. We consequently chose to facilitate a comparison based on Spearman's correlation. We selected Spearman as a non-parametric, rank-based correlation coefficient because no linear relationship between ELISA titers and our positivity ratio can be expected, and the normality assumptions required for parametric tests might be violated. To not bias our correlations by the substantially higher number of negative samples, we draw a random subset of negative samples to exactly match the number of positive samples for the given antibody and antigen combination (see **Fig. 2c**). In **Table S7**, we report the correlation mean and standard deviation for 1000 repetitions of this sampling strategy.

#### QC and visualization of the results

Quality control (QC) statistics give valuable information about the technical issues within the plate quality, variability, and striping due to the dispensing errors as well as edge effects. It also helps in understanding the performance and behavior of positive and negative controls as well as samples[11]. Here, we also monitored the sample transfer by adding nuclear stain to the samples being transferred with an acoustic dispenser. Quality control data analysis pipeline was established to follow the performance of the assay and how different variable parameters behave (**Fig. S4**). This included cell amount/well, and the intensity of DAPI staining, automatically represented as plate layout heatmaps (false color images). The positive and negative controls were plotted for each assay plate (**Fig. S4**), and plate-specific scatter plots were created for each sample/control data point to follow any variation and data distribution. Interactive graphics like scatterplots (**Supp. Data 1**) in html format helps in exploratory data analysis by showing additional information like well annotations and raw values on hovering over the points. For the data visualization per sample, the heatmaps were created for Igs/proteins (**Supp. Data 2** for IgA and IgG).

R programming -based ggplot2, plotly packages and custom functions were used for data processing and visualization[12,13].

**Evaluation of the semi-automated assay results**

Similarly, to the automated version of IFA assay, the images were processed (illumination correction, cell segmentation and feature extraction) with BIAS software. In addition, they were further analyzed in BIAS by supervised machine-learning to predict either positive or negative status of the samples. To determine the positive ratio for the antibodies against SARS-CoV-2 proteins, the number of cells classified as "positive" were divided by the cell-count of transfected (His-tag expressing) cells; parallel values were averaged, excluding the outliers caused by technical errors. To compare the mini-IFA assay results with those from a commercially available antibody test, we performed a recomWell SARS-CoV-2 IgG ELISA, which uses the N antigen (Mikrogen, Germany).

GraphPad Prism Version 5.03 was used to illustrate the results and perform statistical analyses including correlations with patient data. Unpaired t test with Welch's correction was used to compare two groups showing significantly different variances. Due to the deviation from normality, demonstrated by the Shapiro–Wilk test, correlation analyses were performed by calculating the nonparametric Spearman correlation coefficient and the related p value. In general, p values less than 0.05 were considered statistically significant.

**Enzyme-linked immunosorbent assay (ELISA)**

An in-house ELISA for SARS-CoV-2 R, N, and S antigens was used for generating the reference data for sample sets of all sample donors in Finland (**Table S5**) as described[1,2,14]. In addition to traditional ELISA, described in[1,2,14], and used here for N protein with colorimetric detection, we set up an automated high-throughput ELISA on 384-well plate format as follows: Plates (Nunc 384-well MaxiSorp, non-sterile, #460372 for S and R) were coated with purified antigens[2] in 20 µl 0.05 M carbonate buffer (pH 9.6, Medicago, Sweden) overnight at 4°C, S protein 1 µg/ml, and R antigen 1.5 µg/ml. Dispensing was performed with FritzGyger CertusFlex (0.3 mm nozzle, 0.2 bar for all dispenses). For each 384-well plate with serum samples, six plates were coated with each antigen to give two replicate plates for each detection antibody conjugate (anti-IgG/A/M). Plates were covered with MicroClime lids (Labcyte/BeckmanCoulter) filled with ultra-pure water during all incubations through the assay. After antigen coating, the wells were emptied with BioTek EL406 96 pin washer manifold, and 30 µl blocking buffer (3% non-fat milk in PBS with 0.1% Tween 20) added to all wells with EL406 syringe pump. Blocking solution was incubated for 1h RT, and the wells were washed once with 60 µl of PBST (PBS with 0.05% Tween 20) using EL406. Washing solution (PBST) was dispensed with a syringe pump. For serum sample dilution, 15 µl of assay buffer (1% non-fat milk in PBS with 0.05% Tween 20) was added with CertusFlex. Serum samples were pre-diluted in concentration of 1/25 to assay buffer on 384 intermediate plate (Axygen 384-well Clear V-Bottom 240 µL Polypropylene Deep Well Not Treated Plate, #P-384-240SQ-C) pipetting with Biomek FXp 96 head (BeckmanCoulter). Biomek FXp 384 head was used to pipette 5 µl of 1/25 serum dilution to 15 µl of buffer on assay plates to end up with 20 µl of 1/100 dilution of serum. Serum dilution was incubated for 2 h RT with orbital shaking 600 rpm (Heidolph Titramax 1000). Plates were washed with EL406 washer manifold and syringe pump dispensing three times with 80 µl of PBST and aspirated empty. Horseradish peroxidase (HRP) conjugated secondary antibodies were diluted in an assay buffer as follows: goat anti human IgG-HRP 1/6000, goat anti human IgA-HRP 1/5000, and goat anti human IgM-HRP 1/1500. Antibody solutions were dispensed to 20 µl by CertusFlex. Wells without samples and without antigen coating were filled with an assay buffer only. After incubation of 1 h RT plates were washed three times with 80 µl PBST as previously, and then wells filled with Pierce™ ECL Western Blotting Substrate (#32106) using CertusFlex. After 10 min of incubation, the plates were read for luminescence BMG Pherastar FS, with measurement interval of 1 sec, and focal height 12.5 mm for Nunc plate.

For Hungarian SARS-CoV-2 positive serum samples (Orthosera Ltd., see Table S5), the commercially available recomWell SARS-CoV-2 IgG ELISA (Mikrogen, Germany) with purified recombinant SARS-CoV-2 N protein was performed following the manufacturer’s instructions.

**Data availability statement**

Extracted single cell features to train and test models are available at Zenodo: https://dx.doi.org/10.5281/zenodo.6352550.

**Code availability**

Source code is available at GitHub repository: https://github.com/fimm-covid-19-hca/mini-IFA_paper.

**Supplementary Text File 3. Results of the semi-automated mini-IFA method**

Using the semi-automated method (**Table S2, Fig. S5, Supp. Text File 2 for Methods**), the human IgG and IgA responses and the transfection efficacy (for S, N or R antigens) were investigated side by side in each well on the patient sample set #H1 containing 165 samples from 145 COVID-19 patients (median age 44, IQR 34-53) and #H2 containing 200 negative control sera (**Table S5; Supp. Text File 2**). In set #H1, the SARS-CoV-2 infection was confirmed with PCR and/or nucleocapsid-specific IgG ELISA test. Results of IgM analysis are not discussed here due to the lower specificity and sensitivity in the assay performance.

**Assay performance.** Single-cell level phenotypic analysis was done in this case using all analyzed plates and there was no separate test dataset. Confusion matrices show 10-fold cross-validation results with high accuracy for predictions (all > 92%) (**Fig. S8**). Positive and negative sample groups were separated from each other using a dot blot distribution diagram (**Fig. S9A**). SARS-CoV-2 -infected patients did not show IgG and/or IgA positivity against all the three virus proteins, leading to high distribution ranges of the positive samples (Fig. S9A). The N-protein specific IgG data from our assay correlated strongly with the same data analyzed by ELISA (n=117, r=0.8789, p<0.0001) (**Fig. S9B**).

**Clinical observations.** Using the transfection-controlled mini-IFA protocol, both IgG and IgA responses showed positive correlations with age – as previously observed also by Yang et al.[1] – potentially related with trained immunity[2,3] (**Fig. S9C**). IgG positivity, but not IgA, also showed weak correlations with length of symptomatic period, which may be a consequence of a prolonged virus replication and stronger immune responses (**Fig. S9D**). Furthermore, the ratio of IgG positive cells was significantly different in symptomatic and asymptomatic patients, suggesting disease severity to correlate with higher antibody levels as reported earlier[4,5] (**Fig. S9D**).

**Fig. S1.**

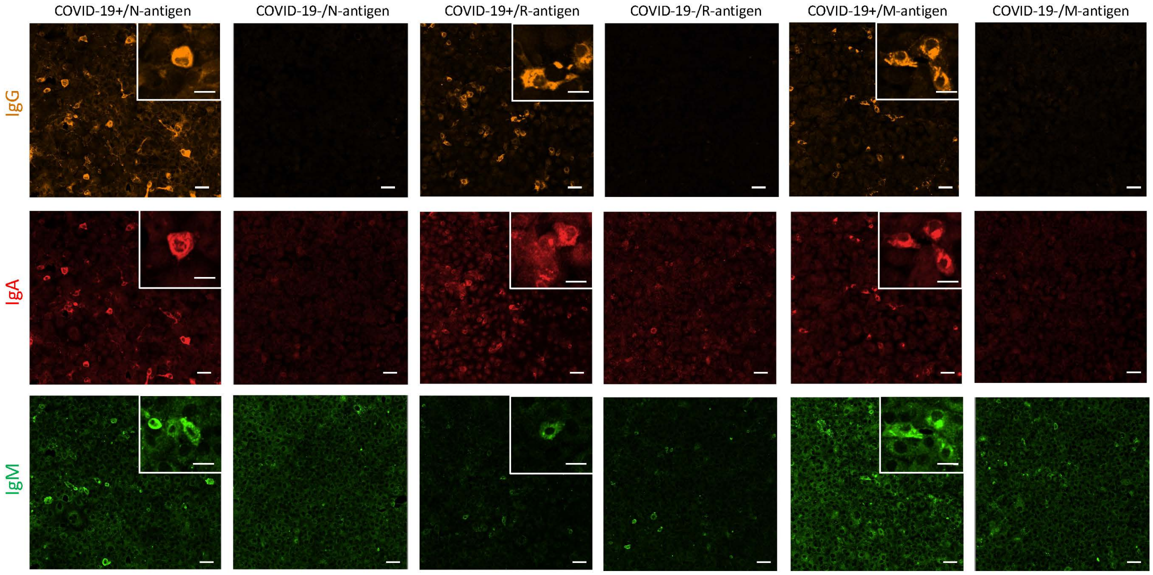


**Fig. S1.** Examples of microscopy images of IgG (DL550, orange), IgA (AF647, red), and IgM (AF488, green) responses to SARS-CoV-2 N, R, and M-antigens, detected from serum samples of COVID-19 patients (COVID-19+) and negative control samples (COVID-19–). (Scale bar: 50 μm in overview images and 20 μm in zoomed images).

**Fig. S2.**

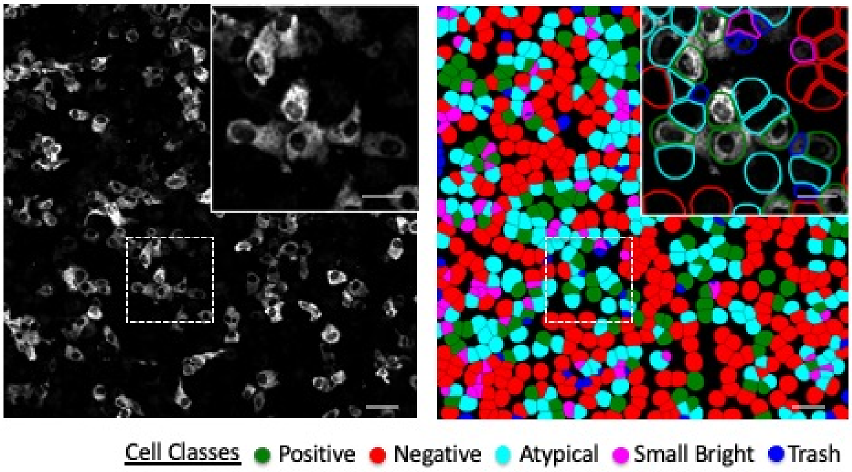


**Fig. S2.** Example of single-cell predictions. Example of the image of S-protein with IgG antibody, scaled linearly from range (quantile(0.5), quantile(0.995)) to 0-255 8-bit values for visualization purposes (left). Single-cell class predictions of the same image made by the model trained for S-protein IgG antibody images (right). The close-up shows the contours of cell boundaries colored with the predicted class color. Scale bar: 50 µm in whole field-of-view images and 25 µm in close-up images. Cells overlapping with antibody signals are classified as Positive, cells without antibody staining are classified as Negative, small, segmented objects are classified as Trash, cells with abnormally small, bright nuclei and a strong intensity of unspecific antibody staining are considered as Small Bright, and some cells next to positive antibody signals are classified as Atypical due to an atypical staining pattern.

Fig. S3a.

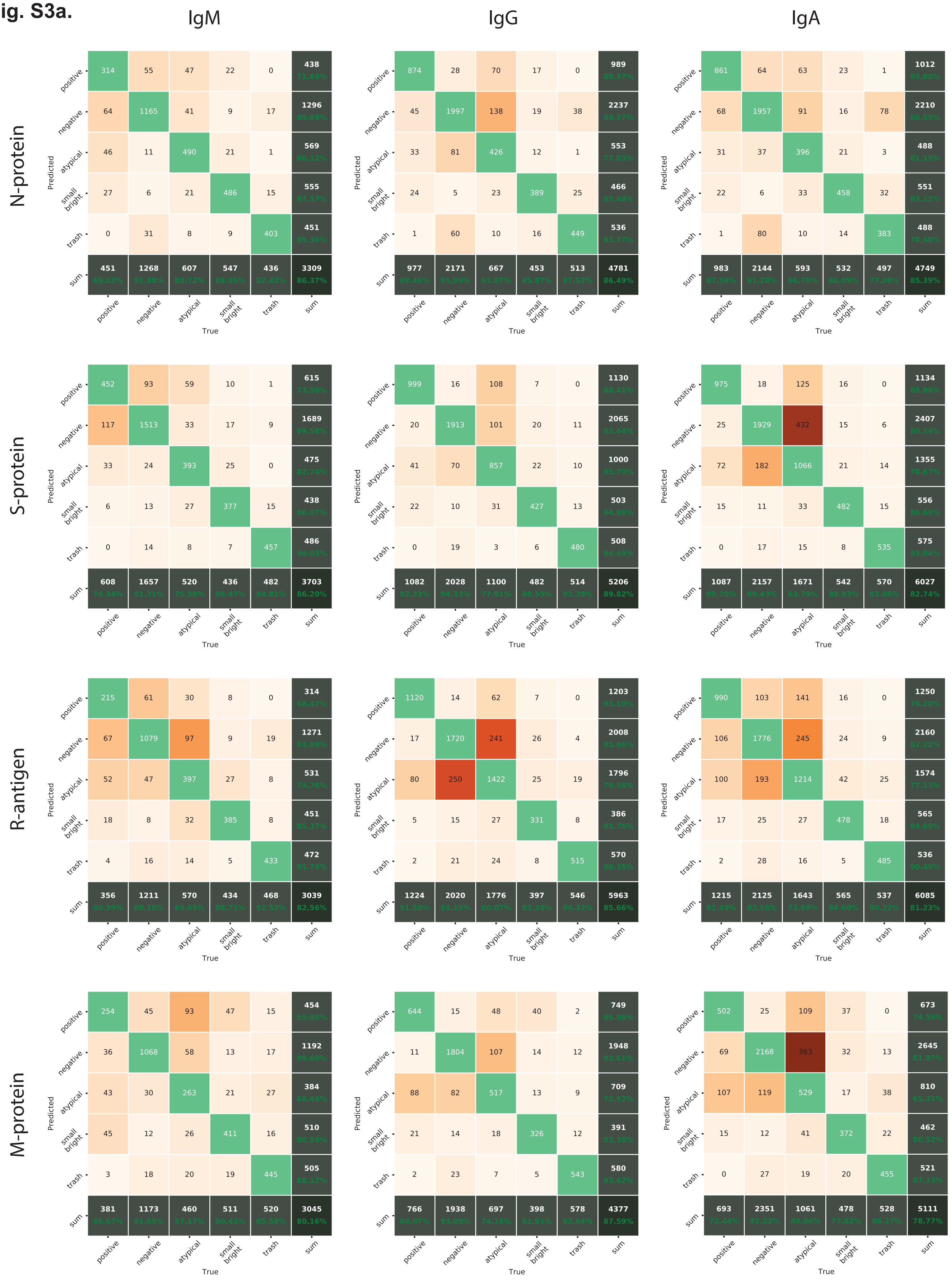
IgM
IgG
IgA
N-protein
S-protein
R-antigen
M-protein
Predicted
True
positive
negative
atypical
small bright
trash
sum
N-protein IgM: positive 314 55 47 22 0 438; negative 64 1165 41 9 17 1296; atypical 46 11 490 21 1 569; small bright 27 6 21 486 15 555; trash 0 31 8 9 403 451; sum 451 1268 607 547 436 3309
N-protein IgG: positive 874 28 70 17 0 989; negative 45 1997 138 19 38 2237; atypical 33 81 426 12 1 553; small bright 24 5 23 389 25 466; trash 1 60 10 16 449 536; sum 977 2171 667 453 513 4781
N-protein IgA: positive 861 64 63 23 1 1012; negative 68 1957 91 16 78 2210; atypical 31 37 396 21 3 488; small bright 22 6 33 458 32 551; trash 1 80 10 14 383 488; sum 983 2144 593 532 497 4749
S-protein IgM: positive 452 93 59 10 1 615; negative 117 1513 33 17 9 1689; atypical 33 24 393 25 0 475; small bright 6 13 27 377 15 438; trash 0 14 8 7 457 486; sum 608 1657 520 436 482 3703
S-protein IgG: positive 999 16 108 7 0 1130; negative 20 1913 101 20 11 2065; atypical 41 70 857 22 10 1000; small bright 22 10 31 427 13 503; trash 0 19 3 6 480 508; sum 1082 2028 1100 482 514 5206
S-protein IgA: positive 975 18 125 16 0 1134; negative 25 1929 432 15 6 2407; atypical 72 182 1066 21 14 1355; small bright 15 11 33 482 15 556; trash 0 17 15 8 535 575; sum 1087 2157 1671 542 570 6027
R-antigen IgM: positive 215 61 30 8 0 314; negative 67 1079 97 9 19 1271; atypical 52 47 397 27 8 531; small bright 18 8 32 385 8 451; trash 4 16 14 5 433 472; sum 356 1211 570 434 468 3039
R-antigen IgG: positive 1120 14 62 7 0 1203; negative 17 1720 241 26 4 2008; atypical 80 250 1422 25 19 1796; small bright 5 15 27 331 8 386; trash 2 21 24 8 515 570; sum 1224 2020 1776 397 546 5963
R-antigen IgA: positive 990 103 141 16 0 1250; negative 106 1776 245 24 9 2160; atypical 100 193 1214 42 25 1574; small bright 17 25 27 478 18 565; trash 2 28 16 5 485 536; sum 1215 2125 1643 565 537 6085
M-protein IgM: positive 254 45 93 47 15 454; negative 36 1068 58 13 17 1192; atypical 43 30 263 21 27 384; small bright 45 12 26 411 16 510; trash 3 18 20 19 445 505; sum 381 1173 460 511 520 3045
M-protein IgG: positive 644 15 48 40 2 749; negative 11 1804 107 14 12 1948; atypical 88 82 517 13 9 709; small bright 21 14 18 326 12 391; trash 2 23 7 5 543 580; sum 766 1938 697 398 578 4377
M-protein IgA: positive 502 25 109 37 0 673; negative 69 2168 363 32 13 2645; atypical 107 119 529 17 38 810; small bright 15 12 41 372 22 462; trash 0 27 19 20 455 521; sum 693 2351 1061 478 528 5111

**Fig. S3b.**

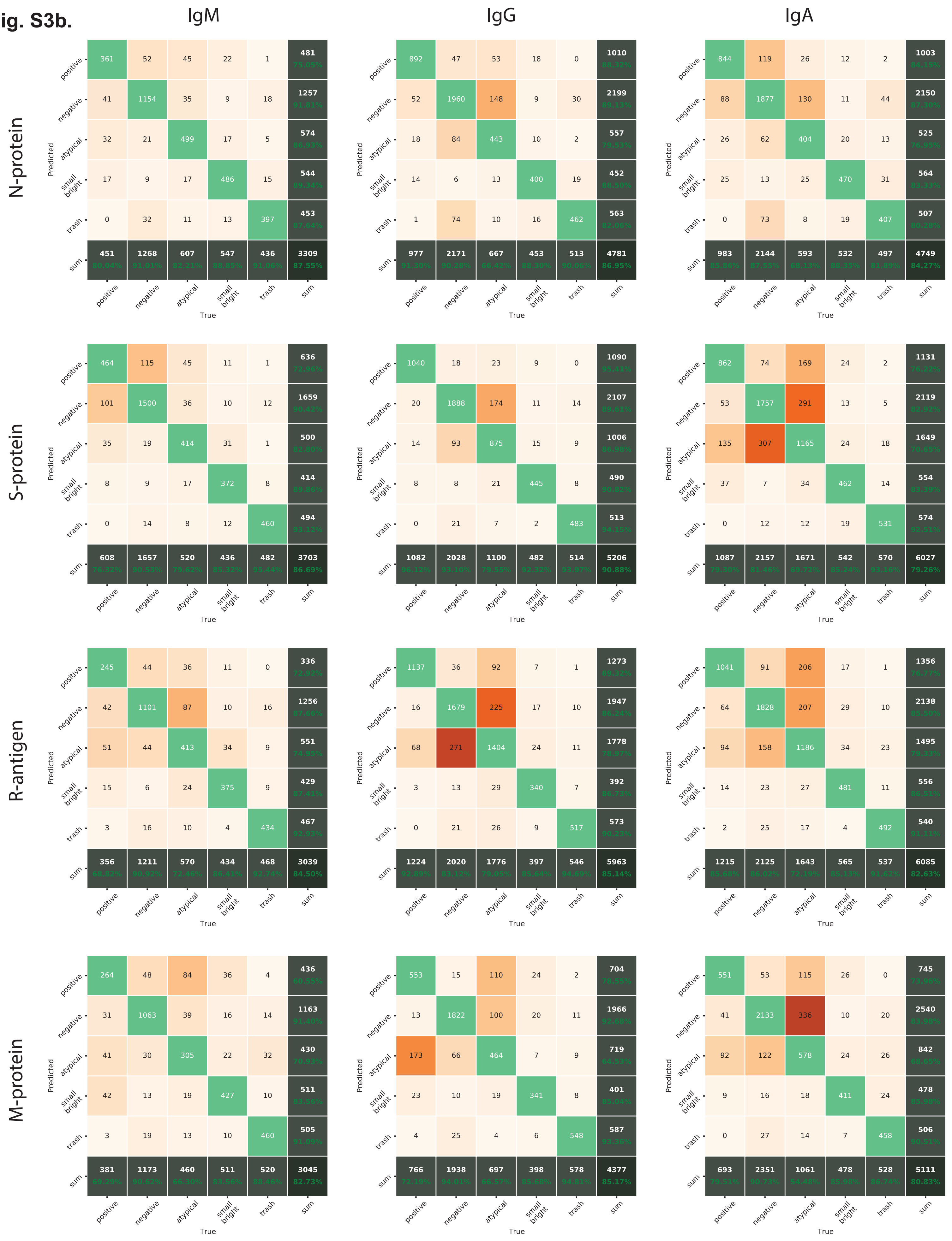


**Fig. S3.** Confusion matrices for the automated method showing the outcome of cross-validation for classifier ANN and each Ig class-antigen pair **a)** with normalization scheme “NegOnly” and **b)** with normalization scheme "PosNegBal".

**Fig. S4.**

a

| Plate | MEAN NUCLEI COUNT | SD NUCLEI COUNT | MEAN DAPI INTENSITY | SD DAPI INTENSITY | MEAN NEG1 IgA | SD NEG1 IgA | MEAN NEG1 IgG | SD NEG1 IgG | MEAN NEG1 IgM | SD NEG1 IgM | MEAN POS1 IgA | SD POS1 IgA | MEAN POS1 IgG | SD POS1 IgG | MEAN POS1 IgM | SD POS1 IgM |
|---|---|---|---|---|---|---|---|---|---|---|---|---|---|---|---|---|
| HX133-09-S01 | 7415,727 | 449,889 | 236,794 | 8,555 | 0,0023 | 0,0017 | 0,0001 | 0,0002 | 0,0044 | 0,0016 | 0,1019 | 0,0211 | 0,1463 | 0,0051 | 0,2749 | 0,0332 |
| HX133-09-S02 | 7104,945 | 337,862 | 186,576 | 4,003 | 0,0031 | 0,0024 | 0 | 0,0001 | 0,0049 | 0,0021 | 0,1325 | 0,0361 | 0,1145 | 0,0204 | 0,3635 | 0,0814 |
| HX133-09-S03 | 6848,531 | 480,027 | 207,890 | 6,984 | 0,0022 | 0,0017 | 0,0001 | 0,0002 | 0,0066 | 0,0043 | 0,1149 | 0,0186 | 0,1468 | 0,0177 | 0,3502 | 0,0643 |
| HX133-09-S04 | 6614,465 | 383,717 | 234,775 | 8,398 | 0,004 | 0,0024 | 0,0001 | 0,0001 | 0,0049 | 0,002 | 0,1644 | 0,015 | 0,2213 | 0,0148 | 0,3781 | 0,0336 |
| HX133-09-N01 | 7469,818 | 287,755 | 244,617 | 12,159 | 0,0001 | 0,0001 | 0,0007 | 0,001 | 0,0088 | 0,0031 | 0,025 | 0,0051 | 0,07 | 0,0194 | 0,3783 | 0,1175 |
| HX133-09-N02 | 7252,073 | 255,875 | 231,867 | 7,483 | 0,0001 | 0,0001 | 0,0022 | 0,0017 | 0,0093 | 0,0026 | 0,0328 | 0,0053 | 0,0725 | 0,0208 | 0,3825 | 0,0811 |
| HX133-09-N03 | 7140,787 | 279,391 | 221,347 | 7,422 | 0,0001 | 0,0002 | 0,0026 | 0,003 | 0,0092 | 0,0047 | 0,0254 | 0,005 | 0,0577 | 0,0085 | 0,3747 | 0,0674 |
| HX133-09-N04 | 7074,227 | 310,412 | 241,163 | 9,224 | 0,0001 | 0,0001 | 0,0028 | 0,002 | 0,0082 | 0,0042 | 0,0393 | 0,0047 | 0,0592 | 0,0126 | 0,4629 | 0,0424 |
| HX133-09-R01 | 7088,907 | 466,600 | 177,954 | 4,227 | 0,0022 | 0,0016 | 0,0004 | 0,0005 | 0,0034 | 0,0013 | 0,0401 | 0,0061 | 0,0699 | 0,0246 | 0,0903 | 0,0551 |
| HX133-09-R02 | 6851,982 | 348,307 | 178,546 | 3,757 | 0,0036 | 0,0014 | 0,0001 | 0,0002 | 0,0032 | 0,0016 | 0,0451 | 0,0159 | 0,0669 | 0,0095 | 0,0719 | 0,0217 |
| HX133-09-R03 | 6657,836 | 396,849 | 194,819 | 6,074 | 0,0023 | 0,0014 | 0,0005 | 0,0005 | 0,0045 | 0,002 | 0,0412 | 0,008 | 0,099 | 0,0054 | 0,166 | 0,0416 |
| HX133-09-R04 | 6337,798 | 487,711 | 234,845 | 8,256 | 0,0043 | 0,0038 | 0,0003 | 0,0004 | 0,0043 | 0,0048 | 0,0358 | 0,0084 | 0,1109 | 0,0101 | 0,0597 | 0,0163 |
| HX133-09-M01 | 7470,417 | 435,013 | 241,035 | 8,696 | 0,0006 | 0,0002 | 0,0007 | 0,0006 | 0,0043 | 0,003 | 0,0486 | 0,009 | 0,0578 | 0,0171 | 0,2523 | 0,0814 |
| HX133-09-M02 | 7229,382 | 403,754 | 232,740 | 6,873 | 0,0004 | 0,0002 | 0,0008 | 0,0007 | 0,0052 | 0,0025 | 0,0582 | 0,0093 | 0,0424 | 0,0125 | 0,0971 | 0,034 |
| HX133-09-M03 | 7372,956 | 363,120 | 219,076 | 6,331 | 0,0002 | 0,0002 | 0,0007 | 0,0005 | 0,0064 | 0,0034 | 0,0599 | 0,0049 | 0,0401 | 0,0094 | 0,1578 | 0,0561 |
| HX133-09-M04 | 7103,130 | 371,408 | 242,631 | 8,777 | 0,0004 | 0,0002 | 0,0015 | 0,001 | 0,0059 | 0,0034 | 0,0791 | 0,0088 | 0,1038 | 0,012 | 0,1713 | 0,0612 |

b

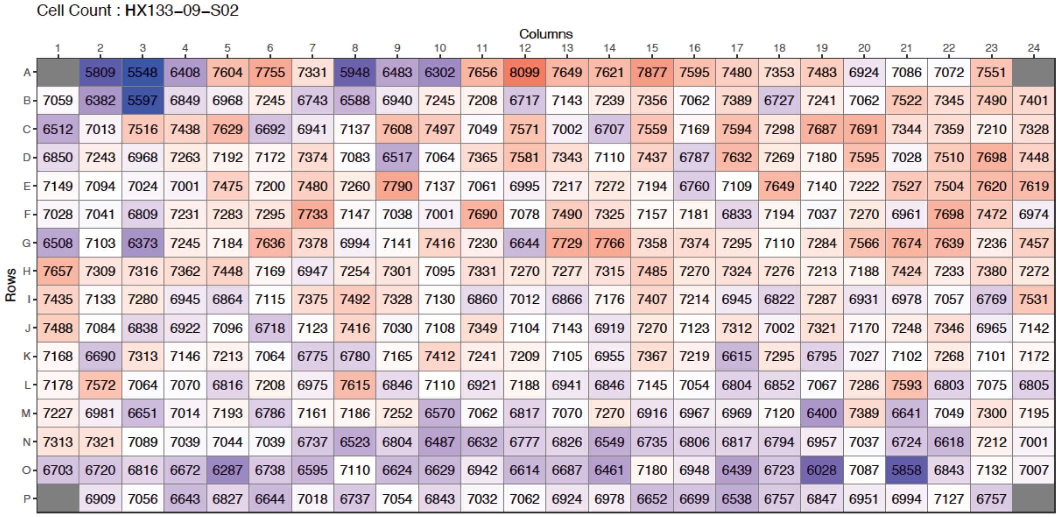


c

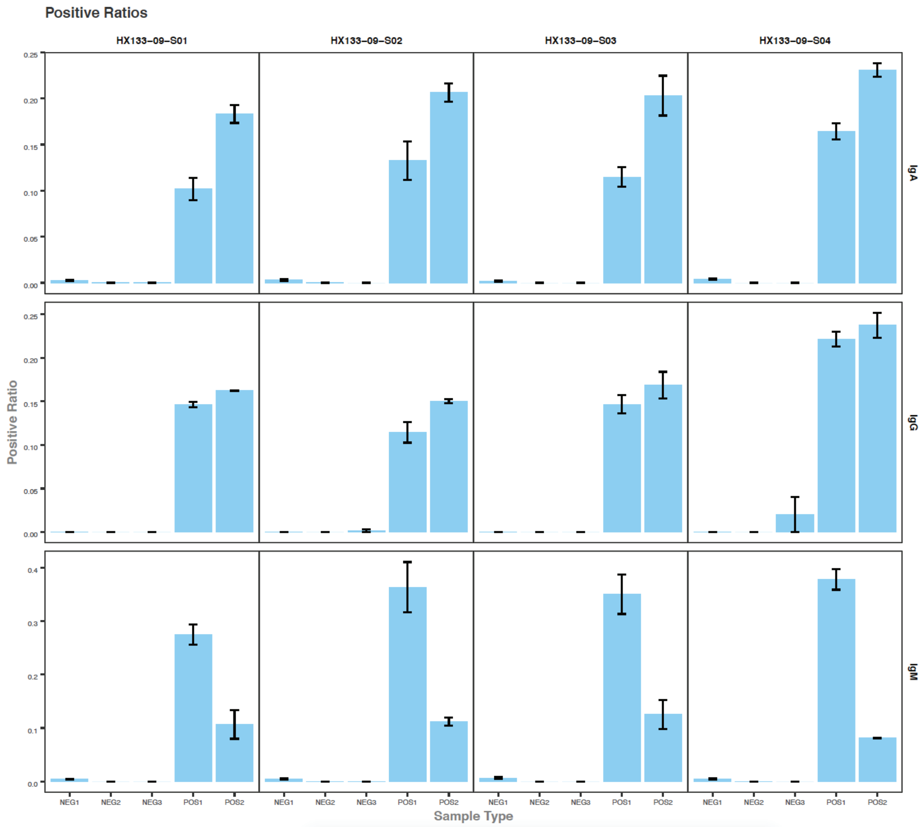


**Fig. S4.** Quality control and data visualization pipeline for the mini-IFA assay (see also Supp. Data 1-2). **a)** The accuracy of the assay was assessed by calculating various parameters across the cells, including nuclei count and staining intensity, as well as by Z prime, and different calculations of values from positive (POS1; COVID-19+ patient sample) and negative (NEG1; COVID-19– control sample) specimens. **b)** Cell count/well across the plates is visualized by heatmaps. **c)** Graphs were created to show the ratio and standard distribution of cells classified as positive (i.e., presence of antibodies against a selected antigen) per total cell count (Y-axis; positive ratio) in the assay control sera (NEG1-3; negative controls; POS1-3; positive controls; X-axis), used for the assay of each antibody/antigen pair.

**Fig. S5.**

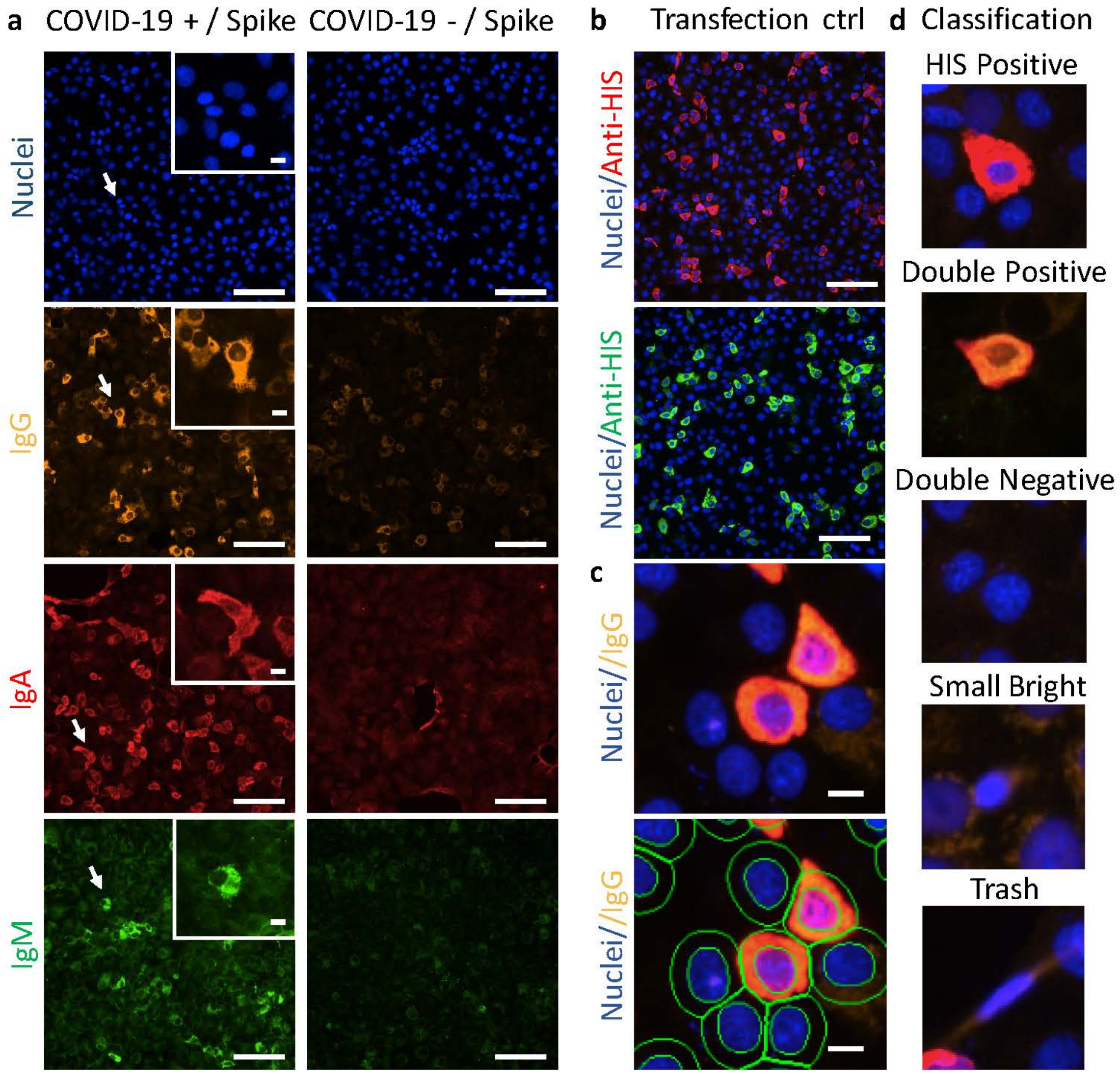


**Fig. S5.** Semi-automated assay pipeline. **a)** Example of microscopy images of IgG, IgA, and IgM labelling on positive and negative control samples. (The scale bar equals to 100 µm in overview images and 10 µm in zoomed images.) **b)** Transfection control staining was used to determine the percentage of transfected cells expressing the viral antigen (co-staining with Igs). The cells positive for both the transfected protein and Igs were annotated as "double positive" cells. Here, example images of HIS-tagged S antigen, recognized either by AF647- (for co-stained IgG/IgM) or AF488- (for co-stained IgA) -conjugated HIS-tag antibodies are shown (scale bars equal to 100 µm). **c)** The steps of evaluation included the segmentation of nuclei first, followed by creating an additional mask for the 'whole cell' by dilating the nucleus area for a maximum of 7 µm, and the 'cytoplasm' was determined by removing the segmented nucleus from the cell (scale bars equal to 10 µm). **d)** For the training set, cells were divided into five classes: HIS positive, double positive, double negative, small bright and trash. All the microscopy images have been taken of cells expressing the S protein, and they have been adjusted from the original 16-bit image format for schematic reasons.

**Fig. S6**

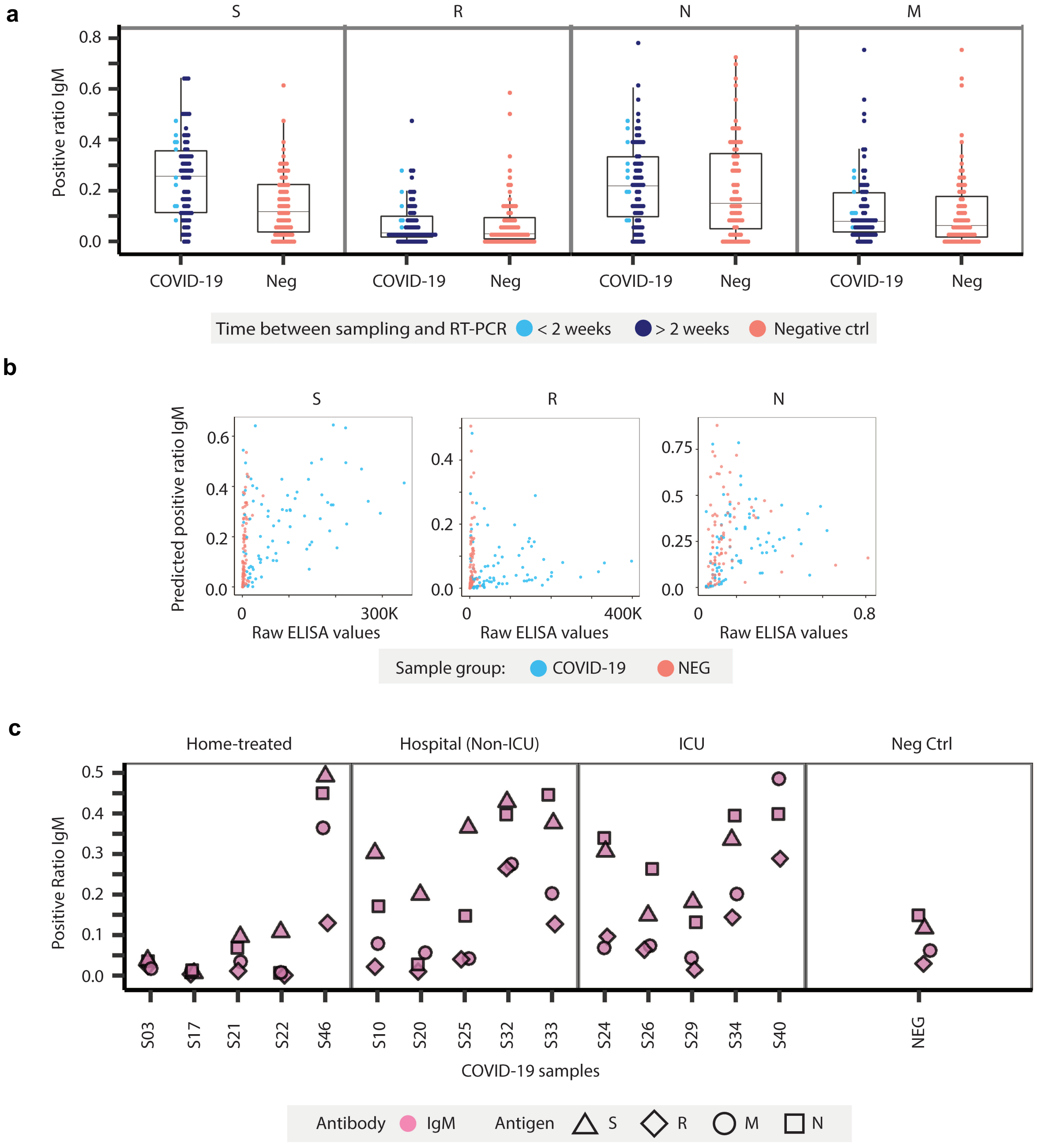


**Fig. S6.** IgM performance in the assay. **a)** The dot plots show the distribution of sample prediction values for IgM responses to the S, R, N, and M antigens in serum samples obtained from COVID-19 positive (COVID-19; collected in 2020) and negative (Neg.; collected in 2017) patients. The serums obtained from COVID-19 positive patients are marked to show the time from the SARS-CoV-2 positive RT-PCR result. See Table S6 for statistics. **b)** Correlation of assay-predicted values with ELISA data for IgM against the S and R antigens (HT-ELISA; n=42 COVID-19 patients, 80 samples; NEG n=80 patients, 80 samples), and against the N antigen (traditional ELISA; n=42 COVID-19 patients, 79 samples; NEG n=80 patients, 80 samples). ELISA values for the S/R vs. N antigen differ in scale as they were obtained via chemiluminescence vs. colorimetric detection, respectively. See Table S7 for statistics. **c)** Patient-specific IgM antibody responses at different time points from the onset of symptoms or from obtaining a positive SARS-CoV-2 RT-PCR result plotted according to COVID-19 severity (treated at home, hospitalized/non-ICU, and hospitalized/ICU).

**Fig. S7**

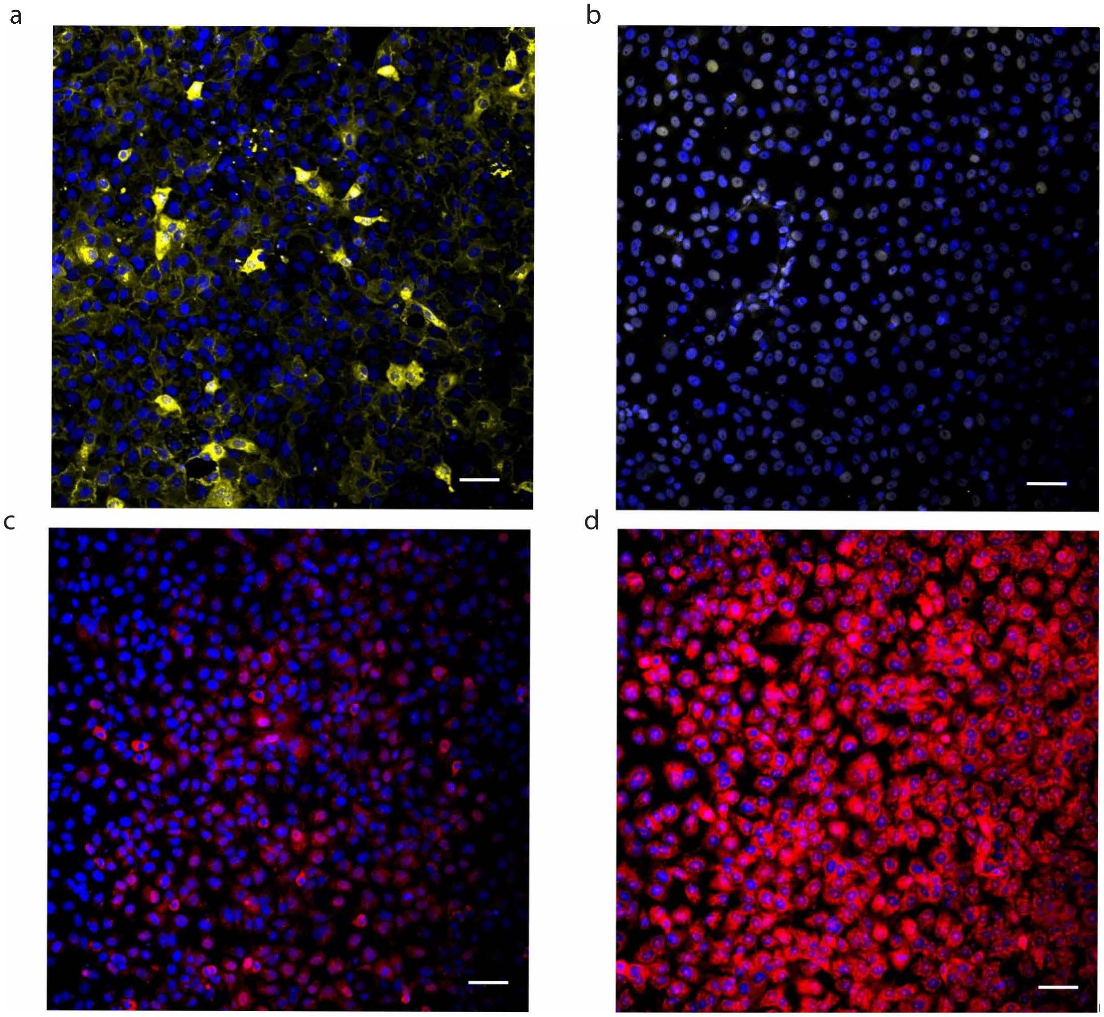


**Fig. S7**. Examples of expert annotation task images. Experts were asked to annotate a total of 576 images as antibody positive or negative. In case they were uncertain, the image was marked as unclear. **a)** Image of IgG antibody/N protein, annotated as positive with perfect consensus by the experts. **b)** Image of IgG antibody/N protein, annotated as negative with perfect consensus by the experts. **c)** Image of IgA antibody/R-antigen with mixed consensus between the experts' opinion. Three experts annotated the image as negative, two as positive, and one as unclear. **d)** Image of IgA antibody/R-antigen annotated as unclear by all the experts. Scale bar: 50 µm.

**Fig. S8.**

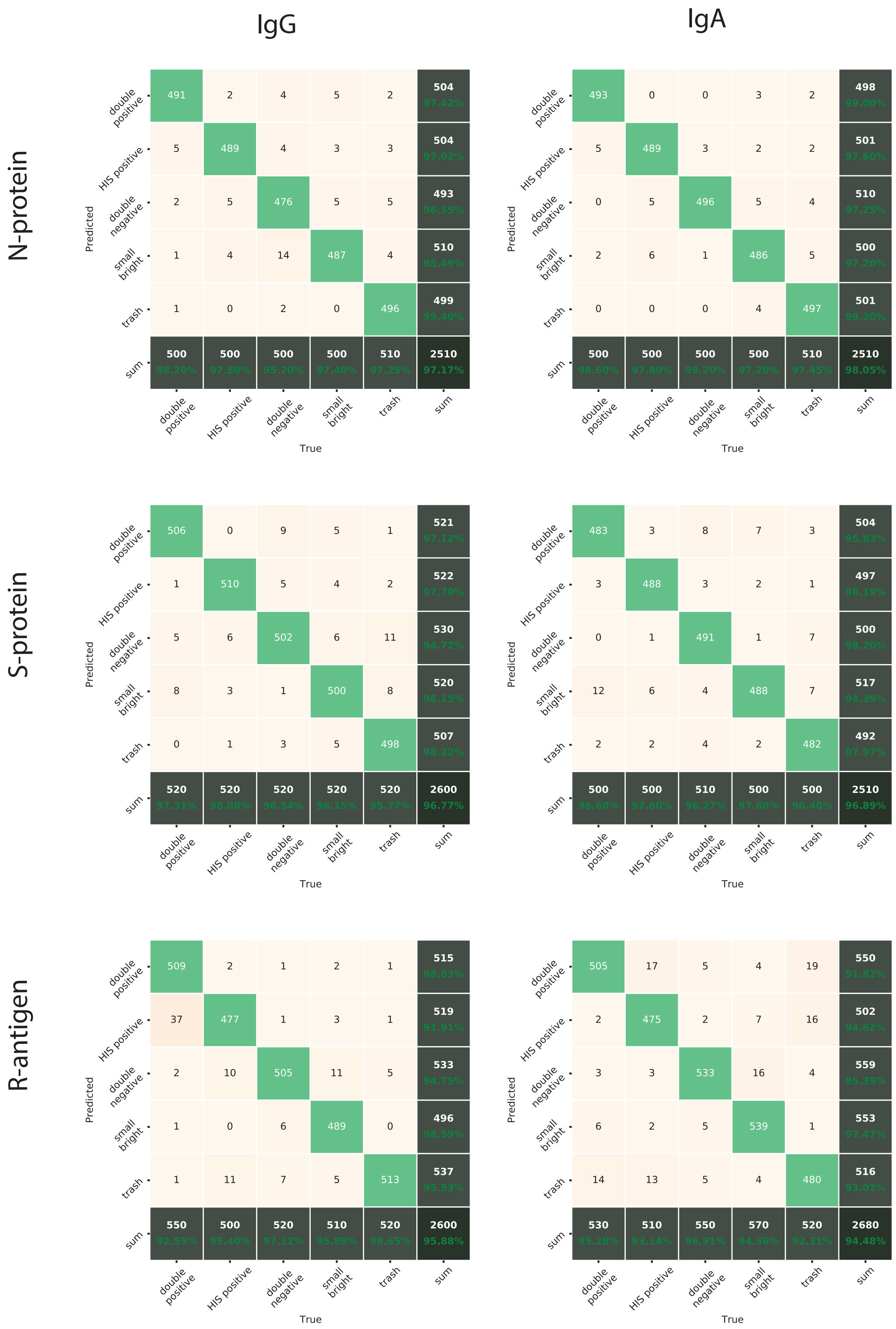


**Fig. S8.** Confusion matrix for the semi-automated method showing the outcome of SVM classifier cross-validation accuracy for IgA/IgG pair with antigen N/S/R.

Fig. S9.

a

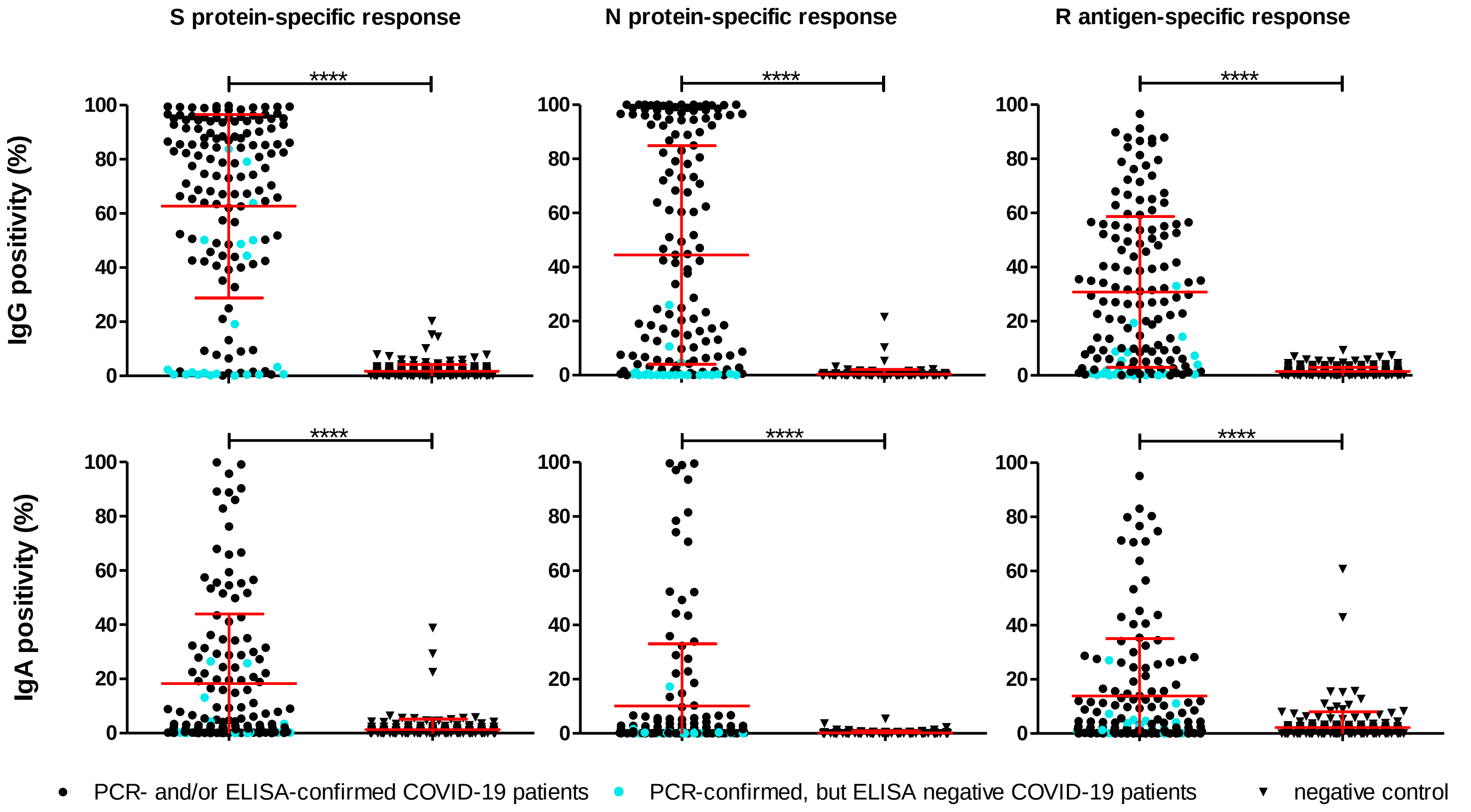
S protein-specific response
N protein-specific response
R antigen-specific response
IgG positivity (%)
IgA positivity (%)
****
0
20
40
60
80
100
PCR- and/or ELISA-confirmed COVID-19 patients
PCR-confirmed, but ELISA negative COVID-19 patients
negative control

b

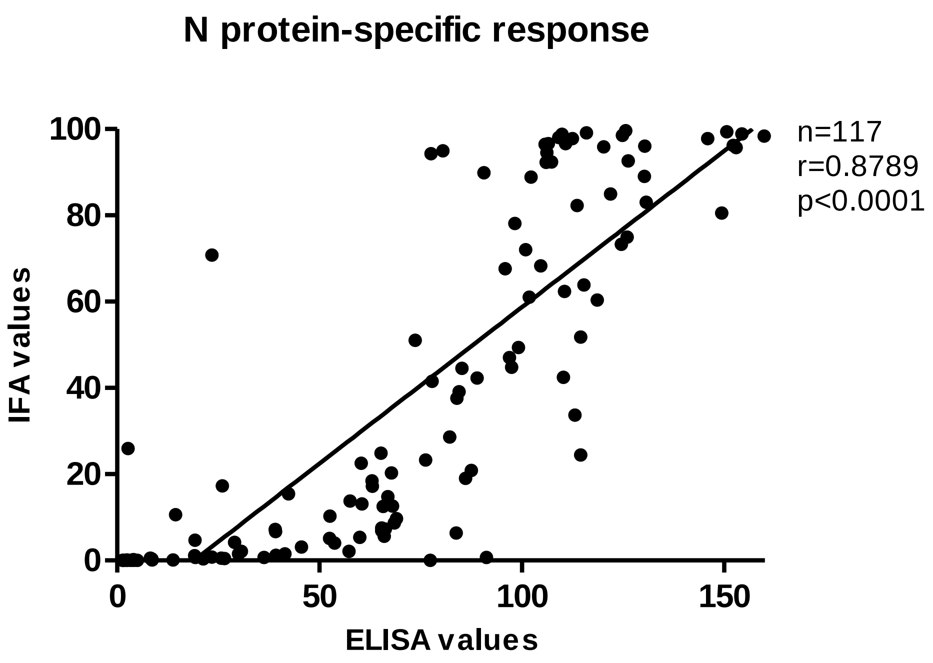
N protein-specific response
n=117
r=0.8789
p<0.0001
IFA values
ELISA values
0
20
40
60
80
100
0
50
100
150

c

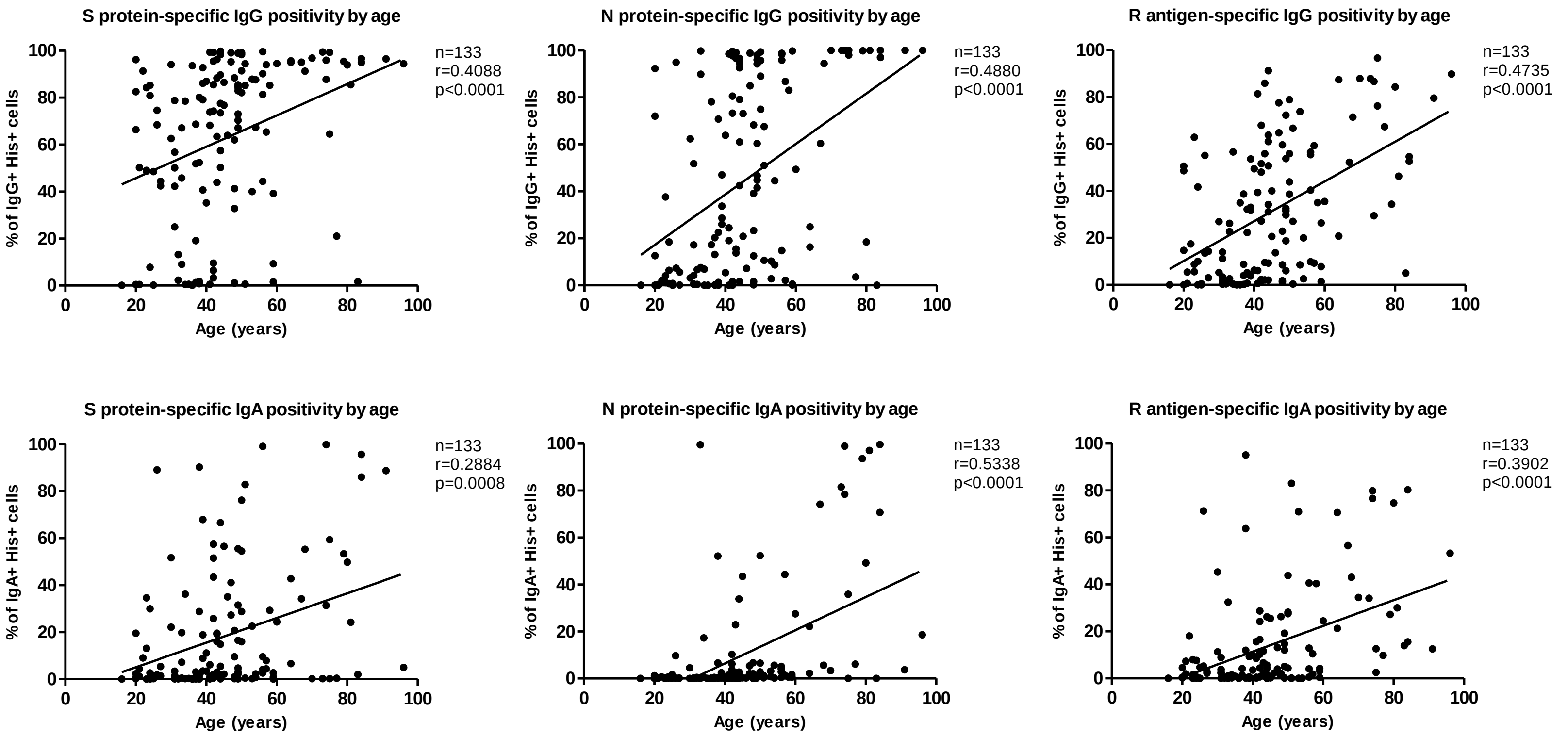
S protein-specific IgG positivity by age
n=133
r=0.4088
p<0.0001
N protein-specific IgG positivity by age
n=133
r=0.4880
p<0.0001
R antigen-specific IgG positivity by age
n=133
r=0.4735
p<0.0001
S protein-specific IgA positivity by age
n=133
r=0.2884
p=0.0008
N protein-specific IgA positivity by age
n=133
r=0.5338
p<0.0001
R antigen-specific IgA positivity by age
n=133
r=0.3902
p<0.0001
% of IgG+ His+ cells
% of IgA+ His+ cells
Age (years)

d

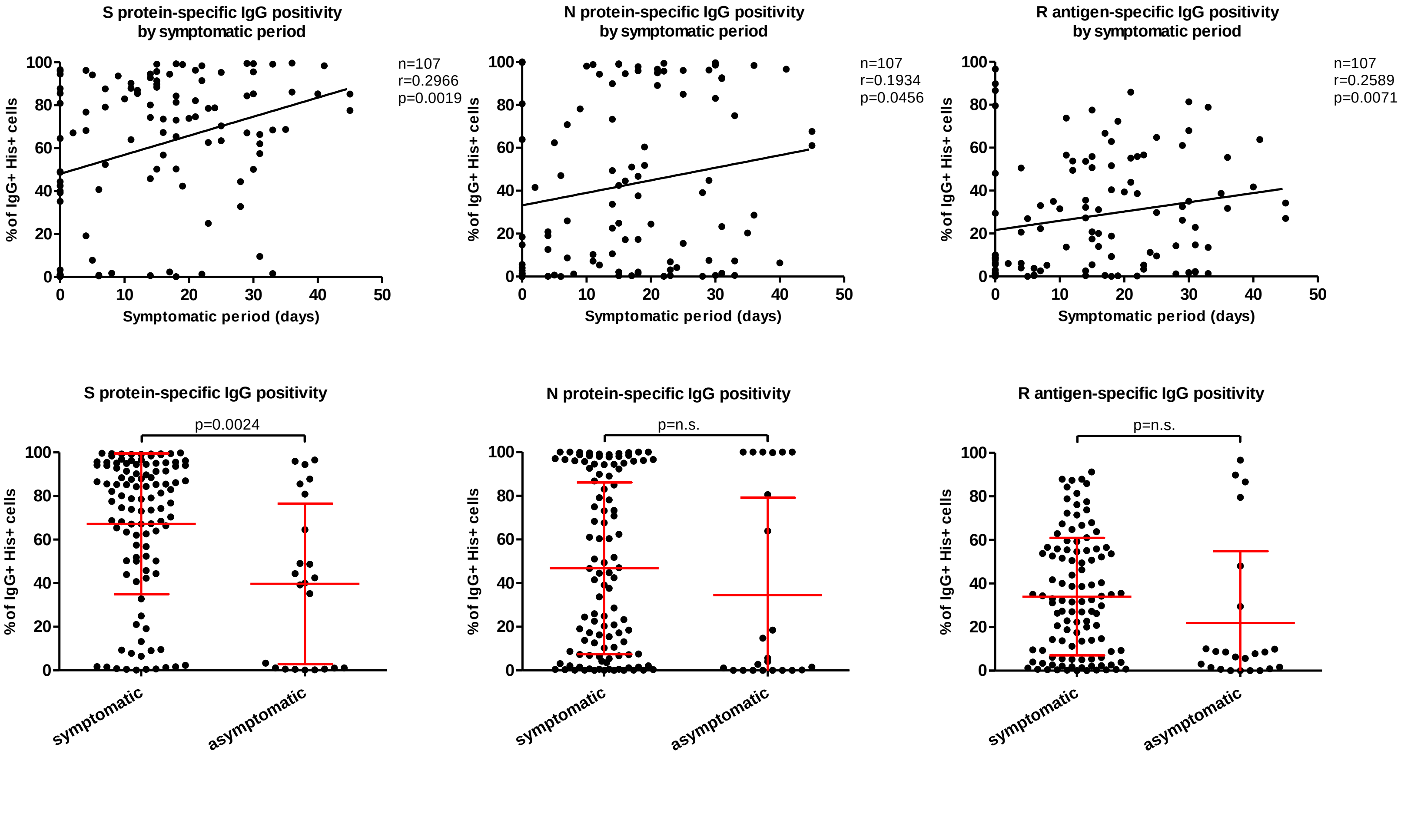
S protein-specific IgG positivity by symptomatic period
n=107
r=0.2966
p=0.0019
N protein-specific IgG positivity by symptomatic period
n=107
r=0.1934
p=0.0456
R antigen-specific IgG positivity by symptomatic period
n=107
r=0.2589
p=0.0071
% of IgG+ His+ cells
Symptomatic period (days)
S protein-specific IgG positivity
p=0.0024
N protein-specific IgG positivity
p=n.s.
R antigen-specific IgG positivity
p=n.s.
symptomatic
asymptomatic

**Fig. S9**. **Performance of the semi-automated version of the mini-IFA assay. a)** Viral protein-expressing cells' positivity for IgG and IgA antibodies. Negative control blood samples were obtained from healthy volunteers (SARS-CoV-2 infection was not investigated; n=200). PCR- and/or ELISA-confirmed COVID-19 patients (n=139) were positive for SARS-CoV-2-specific RT-PCR or nucleocapsid-specific IgG ELISA or both tests (n=96); RT-PCR-confirmed, but ELISA negative COVID-19 patients had positive RT-PCR, but negative ELISA test results (n=21). Average values are mean±SD; **** indicates significance at $p<0.0001$. **b)** Results of the semi-automated assay have shown a strong correlation with ELISA data for nucleocapsid-specific IgG. **c)** Both IgG and IgA antibody responses correlated with patient age (n=133). **d)** IgG positivity correlated with the length of the symptomatic period (n=107). Symptomatic patients (n=99) also showed higher IgG values compared to asymptomatic ones (n=23). Average values are mean±SD (see Supp. Text File 3 for details).

**Supplementary Tables S1-S8**

**Table S1. Materials, reagents, and equipment.** Those used only for semi-automated method are written in cursive.

| **Cell culture** | | | |
|---|---|---|---|
| **Reagent** | **Manufacturer** | **Code** | **Additional info** |
| **Media for Vero E6: MEM + 2mM L-glutamine, + 10% FBS, + 100 IU/mL penicillin and 100 µg/mL streptomycin** | | | |
| MEM: Minimal Essential Medium Eagle | Sigma | M2279-500ml | |
| L-glutamine 200 mM | Gibco® Life Technologies, USA | 25030-024 | |
| Fetal Bovine Serum; FBS | Gibco® Life Technologies, USA | 10270-106 | |
| Penicillin-Streptomycin (5,000 U/mL penicillin, 5000 µg/mL streptomycin) | Gibco® Life Technologies, USA | 15070-063 | |
| Penicillin-Streptomycin (10,000 U/mL) (10,000 U/mL penicillin, 10,000 µg/mL streptomycin) | Gibco® Life Technologies, USA | 15140-122 | |
| 0.25% Trypsin-EDTA | Gibco® Life Technologies, USA | 25200-072 | |
| **Plastic consumables** | | | |
| T-75 cell culture flask | CELLSTAR®, Greiner Bio-one | 658175 | |
| Cellcarrier ultra (PE) 384 imaging plates | PerkinElmer | 6057302 | |
| *T-75 cell culture flask* | *Corning* | *430641U* | |
| *384 Well Microplate, PS, µClear®* | *Greiner Bio-One* | *781090* | |
| *96 Well Microplate, PS, µClear®* | *Greiner Bio-One* | *655090* | |
| **Transfection mix** | | | |
| **Reagent** | **Manufacturer** | | |
| S-pCAGGS, SARS-CoV-2 spike protein/plasmid | Amanat et al. 2020 | - | S antigen |
| RBD-pCAGGS, SARS-CoV-2 receptor-binding domain/plasmid | Amanat et al. 2020 | - | R antigen |
| NP-pCAGGS, SARS-CoV-2 nucleoprotein/plasmid | Rusanen et al. 2021 | - | N antigen |
| M-pEBB, SARS-CoV-2 membrane protein/plasmid | hCoV-19/Finland/1/2020 (Genbank accession MT020781) with pEBB-N-HA mammalian expression plasmid. Synthetized and cloned to the | - | M antigen |

| | | | |
|---|---|---|---|
| | plasmid by GeneArt /Thermo Fisher Scientific | | |
| FuGENE HD | Promega | E2312 | |
| OptiMEM I Reduced Serum Medium | Gibco® Life Technologies, USA | 31985070 | |
| **Staining & Antibodies** | | | |
| **Reagent** | **Manufacturer** | **Code** | **Stock conc.** |
| Rabbit anti-SARS-CoV-2 RBD-His (antiserum) | Rusanen et al. 2021 | | |
| Rabbit anti-SARS-CoV-2 NP-His (antiserum) | Rusanen et al. 2021 | | |
| Goat anti-Human IgG Fc Cross-Adsorbed Secondary Antibody, DyLight 550 | Invitrogen | SA5-10135 | 0.5 mg/mL |
| Goat anti-Human IgM (Heavy chain) Alexa Fluor 488 | Invitrogen | A-21215 | 2 mg/mL |
| Alexa Fluor 647 AffiniPure Goat Anti-Human Serum IgA, α-chain specific | Jackson ImmunoResearch | 109-605-011 | 1.5 mg/ml |
| Mouse monoclonal anti-His-tag antibody, Alexa Fluor 647 | BioLegend | 652513 | 0.5 mg/ml |
| Mouse monoclonal anti-HA-tag antibody 16B12, Alexa Fluor 488 | Invitrogen | A-21287 | 1 mg/mL |
| Goat anti-rabbit IgG(H+L) cross-adsorbed secondary antibody, Alexa Fluor 488 | Invitrogen | A-11008 | 2 mg/mL |
| Hoechst 33342 | Invitrogen | H1399 | 10 mg/mL |
| PBS tablets | Medicago | 09-9400-100 | |
| TBS tablets | Medicago | 09-7500-100 | |
| Tween 20 | Sigma | P9416-100ML | |
| BSA (Bovine Serum Albumin) | Biowest | P6154-100G | |
| Triton X-100 | Sigma | T8787-100ML | |
| Milli-Q $H_2O$ | | | |
| 70% Ethanol | | | |
| *Goat anti-Human IgG (H+L) Cross-adsorbed secondary antibody, Alexa Fluor 546* | *Invitrogen* | *A-21089* | *2 mg/ml* |
| *Mouse monoclonal anti-6x-His Tag antibody (HIS.H8)* | *Invitrogen* | *MA1-21315* | *1 mg/ml* |
| *Goat anti-Mouse IgG (H+L) Highly cross-adsorbed secondary antibody, Alexa Fluor Plus 488* | *Invitrogen* | *A32723* | *2 mg/ml* |

| | | | |
|---|---|---|---|
| *Goat anti-Mouse IgG (H+L) highly cross-adsorbed secondary antibody, Alexa Fluor Plus 647* | *Invitrogen* | *A32728* | *2 mg/ml* |
| *DAPI* | *PanReac AppliChem* | *A1001* | |
| *DPBS (10X), calcium, magnesium* | *Gibco® Life Technologies, USA* | *14080048* | |
| *Pierce™ 20X TBS Buffer* | *Thermo Scientific* | *28358* | |
| *Bovine Serum Albumin* | *Sigma-Aldrich* | *A4503-100G* | |
| *OmniPur Triton X-100 Surfactant - Calbiochem* | *Millipore* | *9410-1L* | |
| **Plastic consumables** | | | |
| Labcyte 384PP (Echo® Qualified 384-Well Polypropylene Source Microplate) | Labcyte | PP-0200 | |
| Costar® 50 mL Reagent Reservoirs | Corning | 4871 | |
| **Equipment** | | | |
| **Use** | **Equipment details** | **Manufacturer** | **other info** |
| Cell seeding | "Brad" BioTek MultiFlo FX dispenser | Biotek | |
| Dispenser(PFA)/washer | "Elmeri" EL406 -plate washer/dispenser | Biotek | |
| Serum dispensing | "Echo" Echo 525 -acoustic dispenser | Labcyte | |
| Dispenser | Certus FLEX 0.3 (air pressure) | Gyger | |
| HCS microscope | Opera Phenix confocal microscope https://www2.helsinki.fi/en/infr bioimaging/fimm-hca | PerkinElmer | |
| Centrifuge for plates | ThermoScientific SL40R | Thermo Fisher | Rotor 75003607 |
| Serum pipetting to Echo source plates | Biomek FXp | BeckmanCoulter | |
| *HCS microscope* | *Operetta CLS High-Content Analysis System* | *PerkinElmer* | |
| *Multichannel pipetting* | *TIPOR-M+ 8 Channel micropipettor* | *Orange Scientific* | |
| *Multichannel pipetting* | *Research plus 12 channel pipette* | *Eppendorf* | |
| **Other components** | | | |
| **Use** | **Equipment details** | **Manufacturer** | **other info** |
| Dispensing cassettes for cells | 5 µl cassette MultiFlo FX | Biotek | |
| Dispensing cassettes for immunostainings | 8 channel 5 µl dispensing EL406 cassette (1260016) | Biotek | |

**Table S2. Transfection efficacy of virus proteins in Vero E6 cells.** Transfection efficiency, shown as transfection % of transfected cells of all cells, of SARS-CoV-2 proteins transfected to Vero E6 cells for HT mini-IFA assay. N, S, and R antigen expression was determined using antibody against HIS-tag (semi-automated method; manual pipetting), or rabbit antibodies produced against N or S/R (automated method). M protein with HA-tag was detected with anti-HA-tag antibody.

| **Automated method (anti-N/S/R/HA-tag ab)** | **N** | **S** | **R** | **M** |
|---|---|---|---|---|
| Average transfection % | 8.67 | 24.96 | 19.43 | 24.73 |
| Median transfection % | 8.74 | 27.83 | 19.59 | 24.73 |
| SE | 0.07 | 0.34 | 0.09 | 0.11 |
| STD | 1.38 | 6.64 | 1.81 | 2.06 |
| n (384-well plate wells) | 384 | 384 | 384 | 384 |
| **Semi-automated method (anti-HIS-tag ab)** | **N** | **S** | **R** | **-** |
| Average transfection % | 18.88 | 18.77 | 27.24 | - |
| Median transfection % | 17.94 | 18.02 | 27.63 | - |
| SE | 0.50 | 0.56 | 0.47 | - |
| STD | 7.76 | 8.64 | 7.33 | - |
| n (96-well plate wells) | 240 | 240 | 240 | - |

**Table S3. Per-cell annotation statistics.** Per-cell Annotation statistics. Per-cell annotations for plates, shown for all expressed virus antigens (N, S, R, and M) and Igs (IgM, IgG and IgA). P=positive, N=negative; A= Atypical; S=Small bright, T= Trash. Dilution indicates the serum dilution used for the analysis.

N-Protein:

| **Run.Plate** | **IgM (dilution 1/25)** | | | | | **IgG (dilution 1/100)** | | | | | **IgA (dilution 1/25)** | | | | |
|---|---|---|---|---|---|---|---|---|---|---|---|---|---|---|---|
| | **P** | **N** | **A** | **S** | **T** | **P** | **N** | **A** | **S** | **T** | **P** | **N** | **A** | **S** | **T** |
| 1.1 | 167 | 282 | 154 | 155 | 105 | 304 | 581 | 125 | 119 | 120 | 263 | 544 | 150 | 155 | 117 |
| 1.3 | 123 | 394 | 118 | 150 | 101 | 270 | 703 | 223 | 101 | 130 | 283 | 864 | 216 | 137 | 110 |
| 1.5 | 102 | 355 | 119 | 142 | 118 | 190 | 692 | 192 | 134 | 159 | 191 | 417 | 72 | 119 | 119 |
| 2.3 | 59 | 237 | 216 | 100 | 112 | 213 | 195 | 127 | 99 | 104 | 246 | 319 | 155 | 212 | 151 |
| **Σ** | **451** | **1268** | **607** | **547** | **436** | **977** | **2171** | **667** | **453** | **513** | **983** | **2144** | **593** | **623** | **497** |
| **Σ** | **3309** | | | | | **4781** | | | | | **4840** | | | | |

S-Protein:

| **Run.Plate** | **IgM (dilution 1/25)** | | | | | **IgG (dilution 1/100)** | | | | | **IgA (dilution 1/25)** | | | | |
|---|---|---|---|---|---|---|---|---|---|---|---|---|---|---|---|
| | **P** | **N** | **A** | **S** | **T** | **P** | **N** | **A** | **S** | **T** | **P** | **N** | **A** | **S** | **T** |
| 1.1 | 179 | 311 | 37 | 113 | 104 | 373 | 836 | 166 | 160 | 140 | 269 | 591 | 401 | 152 | 103 |
| 1.3 | 167 | 398 | 184 | 106 | 105 | 178 | 366 | 329 | 105 | 102 | 232 | 698 | 671 | 136 | 120 |
| 1.5 | 173 | 546 | 117 | 112 | 112 | 245 | 524 | 463 | 122 | 127 | 415 | 509 | 396 | 142 | 123 |
| 2.3 | 89 | 402 | 182 | 105 | 161 | 286 | 302 | 142 | 95 | 145 | 171 | 359 | 203 | 112 | 224 |
| **Σ** | **608** | **1657** | **520** | **436** | **482** | **1082** | **2028** | **1100** | **482** | **514** | **1087** | **2157** | **1671** | **542** | **570** |
| **Σ** | **3703** | | | | | **5206** | | | | | **6027** | | | | |

R-Antigen:

| Run.Plate | IgM (dilution 1/25) | | | | | IgG (dilution 1/100) | | | | | IgA (dilution 1/25) | | | | |
|---|---|---|---|---|---|---|---|---|---|---|---|---|---|---|---|
| | **P** | **N** | **A** | **S** | **T** | **P** | **N** | **A** | **S** | **T** | **P** | **N** | **A** | **S** | **T** |
| 1.1 | 116 | 434 | 106 | 111 | 110 | 239 | 491 | 215 | 108 | 130 | 437 | 540 | 311 | 160 | 158 |
| 1.3 | 84 | 248 | 101 | 132 | 112 | 299 | 618 | 493 | 103 | 134 | 256 | 582 | 589 | 165 | 140 |
| 1.5 | 111 | 292 | 242 | 122 | 134 | 419 | 634 | 828 | 109 | 137 | 386 | 635 | 478 | 118 | 116 |
| 2.3 | 45 | 237 | 121 | 69 | 112 | 267 | 277 | 240 | 77 | 145 | 136 | 368 | 265 | 122 | 123 |
| **Σ** | **356** | **1211** | **570** | **434** | **468** | **1224** | **2020** | **1776** | **397** | **546** | **1215** | **2125** | **1643** | **565** | **537** |
| **Σ** | **3039** | | | | | **5963** | | | | | **6085** | | | | |

M-Protein:

| Run.Plate | IgM (dilution 1/25) | | | | | IgG (dilution 1/100) | | | | | IgA (dilution 1/25) | | | | |
|---|---|---|---|---|---|---|---|---|---|---|---|---|---|---|---|
| | **P** | **N** | **A** | **S** | **T** | **P** | **N** | **A** | **S** | **T** | **P** | **N** | **A** | **S** | **T** |
| 1.1 | 114 | 347 | 159 | 154 | 137 | 121 | 391 | 216 | 135 | 158 | 204 | 453 | 364 | 125 | 112 |
| 1.3 | 107 | 195 | 117 | 132 | 117 | 236 | 704 | 180 | 86 | 192 | 195 | 872 | 177 | 105 | 194 |
| 1.5 | 137 | 444 | 96 | 121 | 123 | 227 | 516 | 135 | 60 | 69 | 156 | 765 | 347 | 130 | 107 |
| 2.3 | 23 | 187 | 88 | 104 | 143 | 182 | 327 | 166 | 117 | 159 | 138 | 261 | 173 | 118 | 115 |
| **Σ** | **381** | **1173** | **460** | **511** | **520** | **766** | **1938** | **697** | **398** | **578** | **693** | **2351** | **1061** | **478** | **528** |
| **Σ** | **3045** | | | | | **4377** | | | | | **5111** | | | | |

**Table S4.** Cross-validation specificity x sensitivity mean and standard deviation to select the best classifier and normalization method.

**IgM**

| RF | Normalisation Scheme | | | | | |
|---|---|---|---|---|---|---|
| | PosNeg | | NegOnly | | PosNegBal | |
| | Mean | StdDev | Mean | StdDev | Mean | StdDev |
| N | 0.71 | 0.14 | 0.66 | 0.2 | 0.72 | 0.08 |
| S | 0.69 | 0.14 | 0.69 | 0.14 | 0.66 | 0.18 |
| R | 0.64 | 0.14 | 0.58 | 0.22 | 0.64 | 0.12 |
| M | 0.67 | 0.04 | 0.65 | 0.07 | 0.64 | 0.07 |
| **Avg** | **0.68** | 0.12 | **0.64** | 0.16 | **0.67** | 0.11 |

| SVM | Normalisation Scheme | | | | | |
|---|---|---|---|---|---|---|
| | PosNeg | | NegOnly | | PosNegBal | |
| | Mean | StdDev | Mean | StdDev | Mean | StdDev |
| N | 0.77 | 0.1 | 0.73 | 0.15 | 0.77 | 0.09 |
| S | 0.77 | 0.11 | 0.75 | 0.13 | 0.75 | 0.13 |
| R | 0.72 | 0.08 | 0.66 | 0.11 | 0.71 | 0.06 |
| M | 0.71 | 0.07 | 0.69 | 0.06 | 0.68 | 0.05 |
| **Avg** | **0.74** | 0.09 | **0.71** | 0.11 | **0.73** | 0.09 |

| ANN | Normalisation Scheme | | | | | |
|---|---|---|---|---|---|---|
| | PosNeg | | NegOnly | | PosNegBal | |
| | Mean | StdDev | Mean | StdDev | Mean | StdDev |
| N | 0.73 | 0.12 | 0.69 | 0.19 | 0.77 | 0.08 |
| S | 0.75 | 0.12 | 0.72 | 0.19 | 0.74 | 0.13 |
| R | 0.64 | 0.1 | 0.61 | 0.14 | 0.68 | 0.09 |
| M | 0.72 | 0.05 | 0.65 | 0.08 | 0.67 | 0.1 |
| **Avg** | **0.71** | 0.1 | **0.67** | 0.15 | **0.72** | 0.1 |

**IgG**

| RF | Normalisation Scheme | | | | | |
|---|---|---|---|---|---|---|
| | PosNeg | | NegOnly | | PosNegBal | |
| | Mean | StdDev | Mean | StdDev | Mean | StdDev |
| N | 0.84 | 0.12 | 0.86 | 0.03 | 0.84 | 0.12 |
| S | 0.91 | 0.06 | 0.86 | 0.04 | 0.9 | 0.07 |
| R | 0.86 | 0.09 | 0.84 | 0.06 | 0.86 | 0.09 |
| M | 0.69 | 0.32 | 0.74 | 0.23 | 0.7 | 0.3 |
| **Avg** | **0.82** | 0.15 | **0.82** | 0.09 | **0.83** | 0.14 |

| SVM | Normalisation Scheme | | | | | |
|---|---|---|---|---|---|---|
| | PosNeg | | NegOnly | | PosNegBal | |
| | Mean | StdDev | Mean | StdDev | Mean | StdDev |
| N | 0.86 | 0.11 | 0.86 | 0.02 | 0.89 | 0.07 |
| S | 0.92 | 0.03 | 0.88 | 0.06 | 0.92 | 0.04 |
| R | 0.89 | 0.06 | 0.91 | 0.01 | 0.89 | 0.03 |
| M | 0.68 | 0.34 | 0.81 | 0.12 | 0.7 | 0.33 |
| **Avg** | **0.84** | 0.14 | **0.86** | 0.05 | **0.85** | 0.12 |

| ANN | Normalisation Scheme | | | | | |
|---|---|---|---|---|---|---|
| | PosNeg | | NegOnly | | PosNegBal | |
| | Mean | StdDev | Mean | StdDev | Mean | StdDev |
| N | 0.87 | 0.09 | 0.87 | 0.02 | 0.89 | 0.06 |
| S | 0.94 | 0.03 | 0.88 | 0.06 | 0.94 | 0.04 |
| R | 0.9 | 0.04 | 0.9 | 0.03 | 0.9 | 0.03 |
| M | 0.69 | 0.33 | 0.82 | 0.11 | 0.72 | 0.33 |
| **Avg** | **0.85** | 0.12 | **0.87** | 0.05 | **0.86** | 0.11 |

**IgA**

| RF | Normalisation Scheme | | | | | |
|---|---|---|---|---|---|---|
| | PosNeg | | NegOnly | | PosNegBal | |
| | Mean | StdDev | Mean | StdDev | Mean | StdDev |
| N | 0.78 | 0.15 | 0.85 | 0.11 | 0.79 | 0.14 |
| S | 0.78 | 0.16 | 0.78 | 0.17 | 0.78 | 0.12 |
| R | 0.72 | 0.14 | 0.67 | 0.17 | 0.75 | 0.15 |
| M | 0.71 | 0.15 | 0.64 | 0.2 | 0.67 | 0.14 |
| **Avg** | **0.74** | 0.15 | **0.73** | 0.16 | **0.75** | 0.14 |

| SVM | Normalisation Scheme | | | | | |
|---|---|---|---|---|---|---|
| | PosNeg | | NegOnly | | PosNegBal | |
| | Mean | StdDev | Mean | StdDev | Mean | StdDev |
| N | 0.79 | 0.15 | 0.8 | 0.12 | 0.79 | 0.14 |
| S | 0.83 | 0.07 | 0.84 | 0.09 | 0.77 | 0.1 |
| R | 0.78 | 0.13 | 0.75 | 0.18 | 0.79 | 0.11 |
| M | 0.72 | 0.12 | 0.7 | 0.12 | 0.73 | 0.09 |
| **Avg** | **0.78** | 0.12 | **0.77** | 0.13 | **0.77** | 0.11 |

| ANN | Normalisation Scheme | | | | | |
|---|---|---|---|---|---|---|
| | PosNeg | | NegOnly | | PosNegBal | |
| | Mean | StdDev | Mean | StdDev | Mean | StdDev |
| N | 0.82 | 0.1 | 0.85 | 0.11 | 0.83 | 0.09 |
| S | 0.84 | 0.08 | 0.86 | 0.08 | 0.78 | 0.11 |
| R | 0.8 | 0.1 | 0.79 | 0.13 | 0.82 | 0.07 |
| M | 0.74 | 0.17 | 0.71 | 0.18 | 0.76 | 0.08 |
| **Avg** | **0.8** | 0.11 | **0.8** | 0.13 | **0.8** | 0.09 |

**Table S5.** The detailed info of the human blood samples used in the study.

| Sample set | Donors (annotations) | Donors (n) | Number of samples (n) | Biospecimen type | Year of collection | Biobanking institution | Method(s) of SARS-CoV-2 diagnosis & serology | Diagnosis | Covid-19 symptoms/clinical status known |
|---|---|---|---|---|---|---|---|---|---|
| #F1a | Sample2020_003 - Sample2020_023 | 21 | 35 | serum | 2020 | Helsinki University Hospital, HUS, Helsinki, Finland | RT-PCR and ELISA, traditional IFA, neutralization assays | COVID-19; 20/21: PCR+ for SARS-CoV-2 | 2/21: hospitalized (non-ICU)/18/21 mild/home-treated |
| #F1b | Sample2020_024 - Sample2020_052 | 24 | 48 | serum, plasma | 2020 | Helsinki University Hospital, HUS, Helsinki, Finland | RT-PCR and ELISA, traditional IFA, neutralization assays | COVID-19; 22/24 PCR+ for SARS-CoV-2 | 8/24: ICU; 10/24 hospitalized (non-ICU) |
| #F2 | Sample2017_001-Sample2017_500 | 500 | 500 | serum | 2017 | Helsinki University Hospital, HUS, Helsinki, Finland | ELISA | negative controls (PUUMALA virus suspected samples) | NA |
| #H1 | AAA0001- AAA514 | 145 | 165 | serum | 2020 | OrthoSera Ltd, University of Szeged, Hungary | RT-PCR and/or ELISA | COVID-19; 139/145 confirmed | mostly but not all (see 139/145 confirmed) |
| #H2 | | 200 | 200 | serum | 2020 | Hungarian National Blood Transfusion Service, University of Szeged, Hungary | - | negative controls | no |

**Table S6**. Statistical values for all Ig classes and viral antigens relating to the comparisons between samples obtained from COVID-19 positive patients (confirmed SARS-CoV-2 infection by RT-PCR; #F1a; b) and COVID-19 negative patients (2017; #F2), related to the plot in Fig. 2a and Fig. S6a.

| Antibody | Antigen | P value | Mean Negative samples | Mean COVID-19 samples | SD Negative samples | SD COVID-19 samples | StdErr Negative samples | StdErr COVID-19 samples |
|---|---|---|---|---|---|---|---|---|
| IgA | M | 1.32603E-07 | 0.01235 | 0.03021 | 0.04156 | 0.04104 | 0.0048 | 0.00474 |
| IgA | N | 3.09893E-10 | 0.00465 | 0.01934 | 0.02253 | 0.02327 | 0.0026 | 0.00269 |
| IgA | R | 2.83964E-03 | 0.02477 | 0.03554 | 0.04436 | 0.05284 | 0.00512 | 0.0061 |
| IgA | S | 8.16885E-11 | 0.03142 | 0.11048 | 0.0659 | 0.09573 | 0.00761 | 0.01105 |
| IgG | M | 2.27817E-18 | 0.0009 | 0.01252 | 0.00148 | 0.01431 | 0.00017 | 0.00165 |
| IgG | N | 1.54312E-24 | 0.00059 | 0.05013 | 0.00191 | 0.0389 | 0.00022 | 0.00449 |
| IgG | R | 6.36239E-21 | 0.00026 | 0.04769 | 0.00053 | 0.03308 | 0.00006 | 0.00382 |
| IgG | S | 1.93984E-22 | 0.00045 | 0.12691 | 0.00209 | 0.05999 | 0.00024 | 0.00693 |
| IgM | M | 2.42417E-01 | 0.12269 | 0.13134 | 0.15213 | 0.14075 | 0.01757 | 0.01625 |
| IgM | N | 1.48921E-01 | 0.20552 | 0.23275 | 0.18753 | 0.16531 | 0.02165 | 0.01909 |
| IgM | R | 2.90875E-01 | 0.07442 | 0.07375 | 0.10824 | 0.08601 | 0.0125 | 0.00993 |
| IgM | S | 1.65103E-05 | 0.14414 | 0.25201 | 0.12584 | 0.161 | 0.01453 | 0.01859 |

**Table S7.** Spearman correlation between predicted positive ratios and ELISA results. Mean correlation and standard deviation are reported for 1,000 repetitions of selecting the same number of positive and negative samples at random.

| | IgM | | IgG | | IgA | |
|---|---|---|---|---|---|---|
| | Mean | StdDev | Mean | StdDev | Mean | StdDev |
| N | 0.37 | 0.02 | 0.82 | 0.01 | 0.7 | 0.02 |
| S | 0.44 | 0.04 | 0.81 | 0.01 | 0.8 | 0.02 |
| R | 0.17 | 0.05 | 0.81 | 0.02 | 0.56 | 0.04 |

**Table S8.** The optimized hyperparameters used for the predictive models.

| Model | | Hyperparameters | |
|---|---|---|---|
| **Protein/Antigen** | **Antibody** | **alpha** | **hidden_layer_sizes** |
| N | IgG | 1.000 | (64) |
| N | IgA | 1.000 | (128,64) |
| N | IgM | 0.050 | (32) |
| S | IgG | 0.100 | (32,32) |
| S | IgA | 1.000 | (32) |
| S | IgM | 0.001 | (32) |
| R | IgG | 0.100 | (32) |
| R | IgA | 0.100 | (32) |
| R | IgM | 1.000 | (128,64) |
| M | IgG | 0.050 | (128) |
| M | IgA | 1.000 | (32) |
| M | IgM | 1.000 | (32,32) |
