## Supplementary measurement data for "Image-based & machine learning-guided multiplexed serology test for SARS-CoV-2"

**Supplementary Data 2**. The heatmap of results implicates the positive cell ratios for IgA/IgG & antigen M/N/R/S pairs in each serum sample.

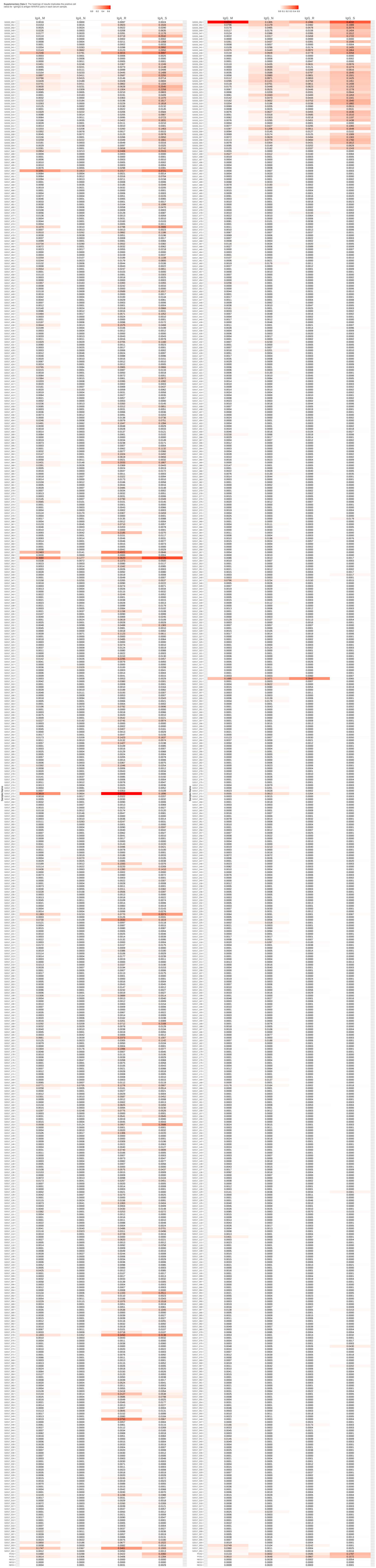
